# A multimodal neural signature of face processing in autism within the fusiform gyrus

**DOI:** 10.1101/2024.01.04.23300134

**Authors:** Dorothea L. Floris, Alberto Llera, Mariam Zabihi, Carolin Moessnang, Emily J.H. Jones, Luke Mason, Rianne Haartsen, Nathalie E. Holz, Ting Mei, Camille Elleaume, Bruno Hebling Vieira, Charlotte M. Pretzsch, Natalie Forde, Sarah Baumeister, Flavio Dell’Acqua, Sarah Durston, Tobias Banaschewski, Christine Ecker, Rosemary J. Holt, Simon Baron-Cohen, Thomas Bourgeron, Tony Charman, Eva Loth, Declan G. M. Murphy, Jan K. Buitelaar, Christian F. Beckmann, the EU-AIMS LEAP group, Nicolas Langer

## Abstract

**Background:** Differences in face processing are commonly reported in case/control studies of autism. Their neural correlates have been explored extensively across single neuroimaging modalities within key regions of the face processing network, such as the fusiform gyrus (FFG). Nonetheless, it is poorly understood how different variation(s) in brain anatomy and function *combine* to impact face processing and social functioning. Extracting the shared information across different modalities is essential to better delineate the complex relationship between brain structure and function, leading to a more comprehensive understanding of the mechanisms underlying autism.

**Methods:** Here, we leveraged data from the large multimodal EU-AIMS Longitudinal European Autism Project (LEAP) to study the cross-modal signature of face processing within the FFG across structural magnetic resonance imaging (MRI), resting-state fMRI (rs-fMRI), task-fMRI (based on the Hariri emotional faces task) and electroencephalography (EEG; recorded when observing facial stimuli) in a sample of 99 autistic and 105 non-autistic individuals (NAI) aged 6-30 years. We combined two methodological innovations: (i) normative modelling was employed on each imaging modality separately to derive individual-level deviations from a predicted developmental trajectory and (ii) unimodal deviations were fused through Linked Independent Component (IC) Analysis to simultaneously decompose the imaging data into underlying modes that characterise multi-modal signatures across the cohort. Next, we tested whether ICs significantly differed between autistic and NAI and whether multimodal ICs would outperform unimodal ICs in discriminating autistic individuals from NAI using a support vector machine under 10-fold cross-validation. Finally, we tested the association between multimodal ICs and cognitive, clinical measures of social or non-social functioning in autism using canonical correlation analysis (CCA).

**Results:** In total, 50 independent components were derived. Among these one multimodal IC differed significantly between autistic and NAI (*t*=3.5, *p*_FDR_=0.03). This IC was mostly driven by bilateral rs-fMRI, bilateral structure, right task-fMRI, and left EEG loadings and implicated both face-selective and retinotopic regions of the FFG. Furthermore, multimodal ICs performed significantly better at differentiating autistic from NAI than unimodal ICs (*p*<0.001). Finally, there was a significant multivariate association between multimodal ICs and a set of cognitive and clinical features associated with social functioning (*r*=0.65, *p*_FDR_=0.008); but not with non-social features.

**Discussion:** The FFG appears to be a central region differentially implicated in autistic and NAI across a range of inter-related imaging modalities and category-selective regions in both the left and right hemispheres. Elucidating more integrated, individual-level neural associations of core social functioning in autism will pave the way for further work on identifying more fine-grained stratification, mechanistic and prognostic biomarkers, and the development of more personalised support.

## Introduction

Autism is a lifelong neurodevelopmental condition with a prevalence of 1 in 36 children^1^. Social-communicative differences are among the most prominent features of autistic individuals^2^. Particularly, difficulties with processing social information and faces, such as perceiving and interpreting facial expressions of emotions and other mental states are thought to have a profound impact on their social functioning and daily living skills^3,4^. While non-autistic individuals (NAI) appear to develop highly skilled strategies to discriminate facial cues at a very early age, autistic individuals have been reported to orient on average less to and acquire less expertise with facial expression recognition^5^. This has, for example, been attributed to a diminished social attention^6^ and structural and functional differences in brain regions implicated in face processing^3,5,7^. While individual neuroimaging modalities have separately been used to characterize the neural correlates of face processing, multimodal studies of key regions associated with face processing remain scarce. Illuminating the rich multimodal information shared across different imaging modalities can unravel complex interactions and variations that may only be partially addressed by single modalities^8^. Specifically, elucidating cross-modal links with regards to face processing in autism will be crucial for understanding the biological mechanisms associated with core social difficulties and paving the way for the development of more personalised support.

The fusiform gyrus (FFG) within the human ventral temporal cortex has been identified as a key neural region associated with higher-order processing of visual stimuli, particularly faces^9^. Extensive neuroimaging research has demonstrated that particularly the fusiform face area (FFA) within the right FFG specifically codes for facial stimuli in typically developing individuals^10,11^. These responses entail increased activation during face perception tasks in functional magnetic resonance imaging (fMRI) studies^12–14^ along with evidence from electroencephalography (EEG) studies showing an event-related potential of negative polarity that peaks at around 170ms when facial stimuli are presented^15,16^. The necessity for a thorough examination of the FFG in isolation is warranted by its detailed, functional heterogeneity. More precisely, the FFG exhibits a fine-grained topographical organization with distinct category-selective patches^17–19^ that are differentially specialized for facial recognition (i.e., FFA)^20^, body part discrimination^21^, object features recognition^22^ and even semantic processing^23,24^. Furthermore, face processing is a lateralized cognitive function with right hemisphere dominance across these modalities^25–28^. An exhaustive examination across different neural signatures of this fine-grained local and hemispheric heterogeneity of the FFG – beyond the FFA – has not been conducted in autistic individuals yet. This can offer valuable new insights in the light of reports of atypical functional specialisation in autism^29,30^.

Accumulating evidence suggests that there is atypical neural organization within the FFG in autistic individuals. Functionally, many studies show that the FFG is hypoactive during face processing fMRI tasks^31–33^ and, atypically connected with the amygdala and superior temporal sulcus^34^ and frontal areas^35^ in autism. Furthermore, EEG studies show that the N170 latency is delayed in autistic compared to NAI^36,37^; and we reported that variation in N170 is associated with change in social behaviour over time^37^. Structurally, there are reports of volume increases in right FFG^38^, a reduction in mean FFG neuron density^39^ and reversed leftward asymmetry^40^ in autism. These atypical neural substrates are thought to be functionally relevant in autistic individuals. For example, they have been linked to differences in facial expression recognition^5^ and face memory^41^, adaptive social functioning^37,42^, and social symptom difficulties and severity^34,41,43,44^.

While these individual imaging modalities (i.e., structural MRI, task-fMRI, resting-state fMRI, EEG) converge to show atypical involvement of the FFG in face processing and related social functioning in autism, there is still little research into how these different neural substrates jointly inform fine-grained FFG organisation and social-communicative functioning in autism. Extracting common information from various modalities is crucial in gaining deeper insights into how brain structure and function reciprocally shape each other, and which common aspects of structure and function inform behaviour, cognition, and clinical conditions such as autism. To date, structure-function coupling has predominantly been addressed via univariate approaches where modalities are combined at the statistical or interpretation level^37,45,46^. However, only when employing multivariate multimodal approaches can we identify direct relationships between different neurobiological mechanisms and how they scale relative to each other. Additional benefits include: (i) we can penetrate across different biological spatial and temporal scales of variation leveraging the unique, complementary aspects covered by each individual imaging modality; (ii) we can gain a more comprehensive understanding of different neurobiological expressions that may converge on a common clinical phenotype (such as atypical face processing and social functioning); (iii) there is a large amount of shared variance across different modalities. Efficient modelling of this has been shown to increase robustness to noise^8,47,48^ and sensitivity to detect potentially subtle effects in high-dimensional data that may otherwise be missed in one single modality^47–52^. Accordingly, prior multimodal efforts are promising as they show that combining information from brain structure and function significantly increases accuracy in predictive frameworks^52–56^. Also in autism, a recent study combining different neuroimaging measures of rs-fMRI, diffusion-weighted imaging and structural morphometry specifically showed that rs-connection topographies within the FFG were differentially implicated between autistic and NAI^57^. While such multimodal endeavours are still scarce in autism, this work specifically underscores the important role of the FFG in the neurobiology of autism. Still, the precise nature and a fine-grained topographical characterization of the multimodal neurobiological interactions within the FFG, and their relationship with the broader clinical phenotype related to social functioning in autism remain to be established.

In the present study, our aim was to provide a more comprehensive understanding of the FFG in face processing in autism by elucidating the simultaneous involvement and multivariate interplay of different neural sources. Such analysis requires both large and deeply-phenotyped samples and given scarce availability, especially in clinical populations, this has previously limited its application. Hence, in this study, we leveraged the unique, large-scale and deeply-phenotyped EU-AIMS Longitudinal European Autism Project^58,59^ (LEAP) which is the largest European multi-centre initiative aimed at identifying biomarkers in autism. This dataset provides a rich set of different neuroimaging modalities, and cognitive, clinical measures as well as tasks related to face processing and social and non-social functioning. Differences in facial expression recognition in autistic individuals have been established in this dataset^5^. To further tap into their multimodal neural correlates, we combined two methodological innovations: (i) first, we employed normative modelling^60^ on each imaging modality separately to derive individual-level deviations from a predicted age-related trajectory. Prior research shows that modelling cortical features as deviations from a normative neurodevelopmental trajectory provides more sensitive measure to map multimodal signatures in psychopathology^52^ while also improving predictive performance^61^. (ii) Next, we conducted multi-modal fusion through Linked Independent Component Analysis^49^ across structural MRI, rs-fMRI, task-fMRI and EEG within the right and left FFG to simultaneously decompose the imaging data into underlying modes that characterise multi-modal signatures differentially in autistic and NAI. We further provided a fine-grained characterization of implicated regions shedding light on the topographic organisation within the FFG in autism. We hypothesized that multimodal components would be more sensitive to capturing subtle diagnostic effects cross-modally and would thus outperform unimodal components in discriminating autistic individuals from NAI. Finally, we hypothesized that joint expression across modalities related to the FFG, and face processing would specifically inform social functioning in autism.

## Methods

### Sample characterization

Participants were part of the EU-AIMS/AIMS-2-TRIALS LEAP cohort^58,59^. They underwent comprehensive clinical, cognitive and MRI assessment at one of six collaborating sites. All autistic participants had an existing clinical diagnosis of autism which was confirmed using the combined information of gold-standard diagnostic instruments, the Autism Diagnostic Interview-Revised^62^ (ADI-R) and the Autism Diagnostic Observation Schedule^63^ (ADOS). The study was approved by the respective research ethics committees at each site (IRAS, UK). Informed written consent was obtained from all participants, or—for minors or those unable to give informed consent—from a parent or legal guardian. For further details on diagnostic procedure, study design and exclusion criteria, see Supplemental Information (SI) and our earlier papers^58,59^. The final sample has both complete imaging data across four different imaging modalities that were integrated (described below) and phenotypic information available. This consisted of 99 autistic individuals, and 105 NAI between 7 and 30 years. For details on demographic information, see Table 1.

### Clinical and cognitive measures

We split available autism-associated measures into two sets of feature sets based on the construct they measure 1) *social-communicative features* comprising measures of difficulties with social communication and daily living skills (i.e., ADOS-social affect, ADI-communication, ADI-social, Vineland Adaptive Behavior Scale^64^ with Communication, Daily Living, Socialization subscales), emotional face matching performance (i.e., Hariri faces task^65^), and social sensitivity to complex emotions (i.e., Reading the Mind in the Eyes test^66^ [RMET]) and 2) *non-social features* comprising restricted, repetitive behaviours (RRBs) (i.e., ADOS-RRB, ADI-RRB, the Repetitive Behavior Scale^67^ [RBS-R]), systemizing (i.e., the Systemizing Quotient^68–70^ [SQ]), shape matching performance (i.e., Hariri shapes task, as the control condition to the Hariri emotional faces task) and sensory processing atypicalities (i.e., Short Sensory Profile^71^ [SSP]) (see Supplement and Table S1). To tackle missing clinical data and to not further reduce sample size, we used imputed clinical data^72^, as in previous work with this dataset^56,73^.

### Region of interest: fusiform gyrus

All analyses were restricted to the right and left FFG based on the Harvard-Oxford atlas (HOA) (FMRIB, Oxford, UK) (i.e., anterior and posterior divisions of the temporal fusiform cortex, temporal occipital fusiform cortex and occipital fusiform gyrus). The size of the ROIs was adjusted to have 100% coverage across all individuals for each imaging modality (for details see SI).

### Imaging modalities

For MRI and EEG data acquisition parameters and detailed preprocessing steps, see SI and Table S2 and S3.

#### Structure – voxel-wise grey matter volume

Voxel-based morphometry (VBM) analyses were run using the CAT12 toolbox (https://neuro-jena.github.io/cat//) in SPM12 (Wellcome Department of Imaging Neuroscience, London, UK). T1-weighted images were automatically segmented into grey matter (GM), white matter, and cerebrospinal fluid and affine registered to the MNI template to improve segmentation. All resulting segmented GM maps were then used to generate a study-specific template and registered to MNI space via a high-dimensional, nonlinear diffeomorphic registration algorithm (DARTEL)^74^. A Jacobian modulation step was included using the flow fields to preserve voxel-wise information on local tissue volume. Images were smoothed with a 4 mm full-width half-max (FWHM) isotropic Gaussian kernel. Features for subsequent normative modeling were VBM-derived, voxel-wise GM volumes per individual restricted to the right and left FFG ROIs.

#### Resting-state fMRI – seed-based connectivity

After recombining the three rs-fMRI scan echoes using echo-time weighted averaging, the rs-fMRI data were preprocessed using a standard preprocessing pipeline that included tools from the FMRIB Software Library (FSL version 5.0.6; http://www.fmrib.ox.ac.uk/fsl). Preprocessing included removal of the first five volumes to allow longitudinal magnetization to reach equilibrium, primary head motion correction via realignment to the middle volume (MCFLIRT), grand mean scaling and spatial smoothing with a 6mm FWHM Gaussian kernel. Next, we thoroughly corrected for secondary head-motion related artifacts, by applying ICA-AROMA, an ICA-based method, which automatically detects and removes motion-related components from the data^75^. ICA-AROMA has been demonstrated to remove head motion-related artifacts with high accuracy while preserving signal of interest^75,76^. Finally, we applied nuisance regression to remove signal from white matter and cerebrospinal fluid, and a high-pass filter (0.01 Hz). The rs-fMRI images of each participant were coregistered to the participants’ anatomical images via boundary-based registration implemented in FSL FLIRT^77^. The T1 images of each participant were registered to MNI152 standard space using 12-parameter affine transformation and refined using non-linear registration with FSL FNIRT (10mm warp, 2mm resampling resolution). Finally, we brought all participant-level rs-fMRI images to 2mm MNI152 standard space by applying the rs-fMRI to T1 and T1 to MNI152 transformations. Next, to characterize the fine-grained functional subdivisions within the FFG in the context of emotional face processing, seed-based correlation analysis was performed between the timeseries derived from each individual’s peak activation voxel within the fusiform face area (FFA) and the remaining voxels within the FFG. This FFA-connectivity was the feature for subsequent normative modeling.

#### Task-fMRI – contrast maps

A well-established task was used to probe functional brain responses during emotional face processing^65^. fMRI data analysis followed standard processing routines in SPM12 (http://www.fil.ion.ucl.ac.uk/spm/), including slice-time correction, a two-step realignment procedure, unified segmentation, and normalization to standard stereotactic space as defined by the Montreal Neurological Institute (MNI), and smoothing with an 8mm full-width-at-half-maximum Gaussian Kernel. Task conditions were modelled as boxcar functions that accounted for the presentation of face blocks and shape blocks, respectively and convolved with the canonical hemodynamic response function (HRF) and subjected as predictors to a general linear model (GLM), along with six realignment parameters to account for head motion. During first-level model estimation, data was high-pass filtered with a cut-off of 256s, and a first order autoregressive model was applied. The face matching condition was contrasted to the shape matching condition to identify brain responses reflecting sensitivity to emotional faces. These T-contrast maps per individual (restricted to the right and left FFG ROIs) were the features for subsequent normative modeling.

#### EEG – source reconstruction

Participants were presented with three repeated, upright, or inverted face stimuli, repeated 168 times over four blocks. Here, all face stimuli were included. The following preprocessing steps were carried out: 1) harmonisation of electrode labels to 62-electrode common montage; 2) deviation of horizontal electrooculogram (HEOG) from electrodes AF7/8; 3) generation of variance-based data quality metrics and extraction of impedance values from Brainvision sites; 4) re-reference to FCz. This process resulted in harmonised data in a common EEGLab^78^ format, upon which all subsequent task-specific analyses were performed. Further offline treatment of the data was done using the FieldTrip toolbox^79^. Raw EEG data were band-pass filtered 0.1 to 30 Hz with 2000-ms padding and epoched from −200 to 800 ms after stimulus onset. Artifacts were identified and removed according to criteria detailed in the SI. Next, beamforming-based source level analysis was conducted within the left and right FFG to derive source estimates for several cortical parcels. The principal component across these time series was used as the feature for subsequent normative modeling. For a detailed description of feature extraction for each modality, see the SI.

### Normative modelling

Normative modelling is an emerging statistical technique that allows parsing heterogeneity by charting variation in brain-behaviour mappings relative to a normative range and provides statistical inference at the level of the individual^80^. The term ‘normative’ should not be seen as incompatible with the neurodiversity framework as it simply refers to statistical norms such as growth charts that vary by demographics such as age and gender. Variation from the norm is part of neurodiversity. Here, we trained normative models^60,80,81^ using Bayesian Linear Regression (BLR)^82^ (https://pcntoolkit.readthedocs.io/en/latest) for each brain imaging modality within the right and left FFG ROI independently using age, sex and scanning site as covariates. A B-spline basis expansion of the covariate vector was used to model non-linear effects of age. Normative models were derived in an unbiased manner across the entire sample under 10-fold cross-validation^52,60,83^. This Bayesian approach calculates the probability distribution over all functions that fit the data while specifying a prior over all possible values and relocating probabilities based on evidence (i.e., observed data). As such, it yields unbiased estimates of generalizability and inferences with increasing uncertainty with fewer data. To estimate voxel-wise/time-point-wise deviations for each modality in each individual, we derived normative probability maps (NPM) that quantify the deviation from the normative model summarized in *Z*-scores. These subject-specific Z-score images provide a statistical estimate of how much each individual’s recorded value differs from the predicted value at each voxel/time-point. The accuracy of the normative model was evaluated using the correlation between the recorded and the predicted voxel values (Rho), the mean standardized log-loss (MSLL), standardized mean squared error (SMSE), and the explained variance (EV) (Figure S1) as well as based on the forward models (Figure S3). Furthermore, we compared model performance when modelling age linearly (without a B-spline basis expansion; Figure S2). To assess whether autistic and NAI differed in their extreme deviations based on *unimodal* features, thresholded Z-scores (Z>|2.6|^30,52,84,85^, corresponding to the 99.5th percentile) were compared between the two groups using a two-sample t-test (see SI). Code is available at https://github.com/amarquand/PCNtoolkit.

### Linked Independent Component Analysis

In order to gain more comprehensive insights into cross-modal signatures of face processing, we merged the different individual-level deviations from all imaging modalities (GM volume, FFA-connectivity, T-maps contrasting the faces condition to the shapes condition, and the principal components of source reconstructed time series) using Linked Independent Component Analysis (LICA)^47,49–52,56,86^ (see SI). This is a Bayesian extension of the single modality ICA model which provides an automatic and simultaneous decomposition of the brain features into independent components (ICs) that characterize the inter-subject brain variability. These multiple decompositions share a mixing matrix (i.e., subject course) across individual feature factorizations that reflect the subject contributions to each IC. These subject loadings per IC were later used to investigate the multivariate relationship between the brain phenotypes and clinical measures (see canonical correlation analysis below). Further, each IC also provides a map of spatial or temporal variation per modality and a vector reflecting the relative contribution of each modality to the component. Here, LICA was used to merge the unthresholded Z-deviation maps derived from normative modeling across the four different imaging modalities within the right and left FFG ROIs. Each measure per hemisphere was treated separately (i.e., right structure, left structure, right rs-fMRI, left rs-fMRI, right task-fMRI, left task-fMRI, right EEG, left EEG) resulting in eight input maps (i.e., modalities). Hemispheres were modelled separately given known brain asymmetric differences in autism^30,44,87^ and to study the hemispheric contributions and model the different noise characteristics individually. We estimated 50 independent components based on our sample size and following recommendations described in earlier papers^50–52,56,86^ (i.e., sample size ∼N / 4). To evaluate the robustness of our selected model order (N=50), we re-ran LICA using different dimensional factorizations of subject loadings (N=40 and N=60) and computed correlations among them. LICA code is available at https://github.com/allera/Llera_elife_2019_1/tree/master/matlab_flica_toolbox.

### Group Differences

The subject loadings of all ICs were compared between autistic and NAI using a two-sample t-test. Multiple comparisons were corrected for using the False Discovery Rate (FDR)^88^. ICs showing significant group differences were further characterized by plotting each contributing modality’s spatial map and temporal profile (Z-thresholded at the 95^th^ percentile). To further characterize the most implicated regions within the FFG per modality, we computed the overlap between supra-threshold voxels and a structural (i.e., the Harvard-Oxford atlas, which covers the entire FFG) and a functional (i.e., a probabilistic functional atlas of the occipito-temporal cortex^18^ which covers category-specific FFG patches) atlas (see SI).

### Multimodal components

Next, given the current work’s focus on multimodal neural sources, we tested the hypothesis whether multimodal components performed superior to unimodal components in differentiating autistic individuals from NAI. For this, we calculated a multimodal index (MMI) per independent component to quantify the multimodal nature of modalities in each IC^50^ (for details, see SI). The MMI ranges from 0 (equating to 100% unimodal contribution) to 1 (equating to equal contributions from all modalities). Multimodal components were defined as each single imaging modality (i.e., regardless of hemisphere) not having more than a 90% contribution to each component and an MMI below 0.1 (see Figure S4). Components below this threshold were regarded as unimodal.

### Autism classification

Next, we implemented two support vector machine (SVM) classifiers with a linear kernel – one using unimodal and one using multimodal components as features to test for the added value of multimodal features. The SVM was trained and evaluated using 10-fold cross-validation and class-weighting was used to account for group size imbalance. The area under the receiver operating characteristic curve (AUC) was used as the performance metric to assess the classifier’s discrimination ability. To test for significant differences in AUC between multimodal and unimodal components, we generated a null distribution of AUC differences by shuffling the cross-validated scores 10.000 times and re-evaluating the classifier performance and computed the likelihood of observing the observed AUC difference under the null hypothesis. To test for robustness of results across different multimodal thresholds, we re-ran analyses across different thresholds resulting in slightly varying degrees of multimodality ranging between 85% to 99% of single modality contributions. Given that each threshold resulted in a different number of unimodal vs. multimodal components, we further checked whether results remained stable when forcing uni- and multimodal components to have the same number of features; for this we selected between the top one and 22 most uni- and multimodal ICs.

### Clinical-cognitive associations

To test for the clinical relevance of multimodal ICs, we ran canonical correlation analyses (CCA)^89^ modelling the multivariate relationship between multimodal ICs and cognitive, clinical features related to either social or non-social functioning (described in detail above) in autistic individuals only. CCA is a multivariate approach to simultaneously model two sets of linear projections (based on the brain-related independent components and the cognitive features) to maximize their correlation. The statistical significance of the CCA modes was assessed by a complete permutation inference algorithm proposed by Winkler *et al.*^90^, where both brain and behaviour data were permuted separately across all participants with 10,000 iterations. In total, we ran two separated CCAs testing the multivariate relationship between the brain measures (multimodal ICs) and *a)* social-communicative features related to social functioning and face processing in autism or *b)* non-social features associated with autism. For further details, see the SI.

To visualize the spatial and temporal patterns of each imaging modality associated with each clinical cognitive measure, we computed the correlations between the original imaging data (i.e., the Z-deviation maps) and the canonical imaging variate (V) derived from the CCA^91^. Significance of correlation maps was assessed with 1000 permutations and significant clusters / timepoints were next visualized and further characterized in terms of their functional and anatomical characteristics by computing their overlap with the probabilistic functional atlas of human occipito-temporal visual cortex (VIS-atlas)^18^ of early visual and category-selective regions (see Figure 2h) and the HOA atlas (see Figure 2m). For further details see the SI.

**Figure 1.**
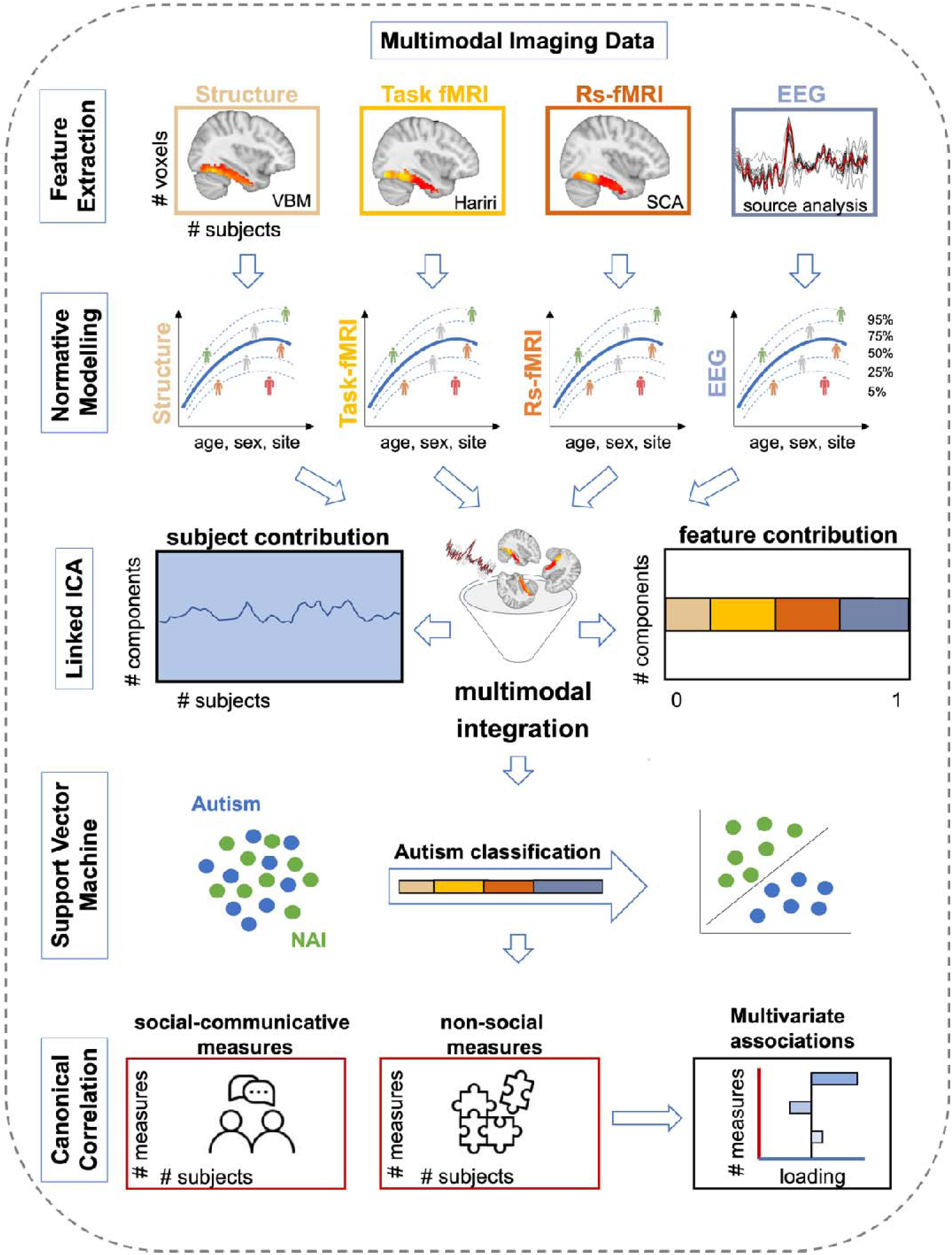
Overview of the methodological approach. Features for each modality were extracted from the right and the left fusiform gyrus. These were: a) grey matter volume based on VBM for structural MRI; b) T-maps contrasting the faces condition to the shapes condition reflecting sensitivity to emotional faces from the Hariri paradigm for task-fMRI; c) seed-based (i.e., fusiform face area) connectivity (SCA) for rs-fMRI; and d) the principal component of different source reconstructed time series for EEG. Next normative modelling was applied to each imaging modality using Bayesian Linear Regression. To model cross-subject individual-level variation, resulting Z-deviation maps per modality were statistically merged using linked independent component analysis resulting in measures of modality contributions and subject loadings. Next, we tested for group differences in ICs and group separability using either multi- or unimodal ICs and compared their performance. Finally, we computed multivariate associations (i.e., canonical correlation analysis) between subject loadings and clinical, cognitive measures related to either social-communicative or non-social features.

**Figure 2.**
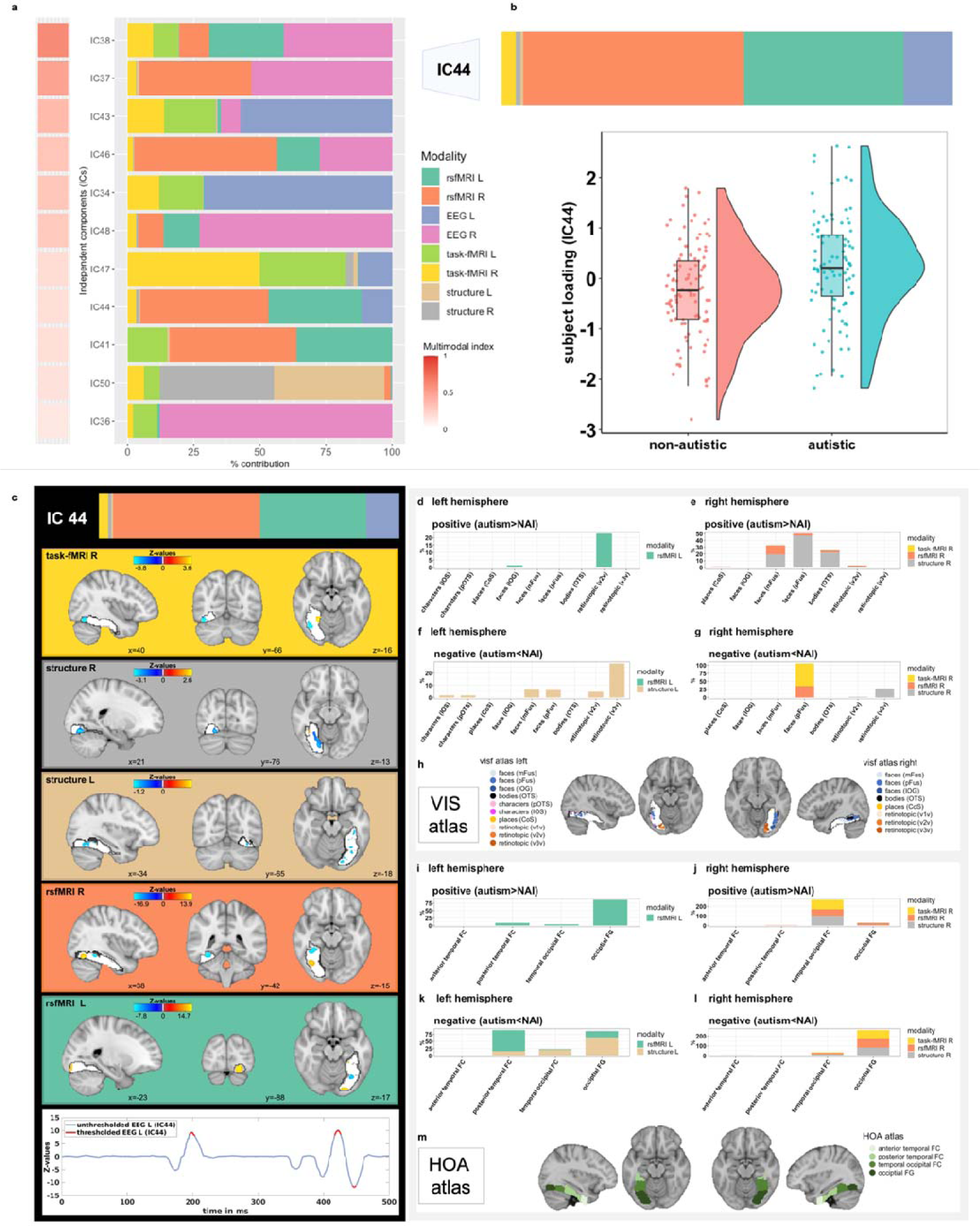
Among all independent components, eleven were considered multimodal (Figure 2a), with a single modality contribution not more than 90%. Among these, IC44 showed a significant group difference where autistic individuals had higher contributions than non-autistic individuals (Figure 2b). Figure 2c shows spatial and temporal Z-maps thresholded at the 95^th^ percentile of the different neuroimaging modalities associated with IC44. Positive values (in yellow) depict positive loadings onto the IC and mean autistic individuals have higher deviations than NAI; whereas negative values (in blue) depict negative loadings onto the IC and mean autistic individuals have lower deviations than NAI. Suprathreshold timepoints are depicted in red. Figures 2d-g depict the spatial overlap of suprathreshold voxels with a probabilistic functional atlas of the occipito-temporal cortex (i.e., VIS-atlas^18^). Figure 2h depicts the VIS -atlas and the different early visual and category-selective subregions covering the FFG. Figures 2i-l show the spatial overlap of suprathreshold voxels with the structural Harvard-Oxford atlas and the four subregions of the fusiform gyrus (i.e., anterior and posterior divisions of the temporal fusiform cortex, temporal occipital fusiform cortex and occipital fusiform gyrus depicted in Figure 2m). Here, Figures 2d-e and 2i-j show the positive loadings (i.e., autism > NAI) and Figures 2f-g and 2k-l the negative loadings (i.e., autism < NAI), whereas Figures 2d/f and 2i/k depict the left hemisphere and Figures 2e/g and 2j/l the right hemisphere.

Finally, in order to assess robustness of CCA results, as previously, we set a range of multimodal thresholds between 85% to 99% and selected components with modality contributions exceeding this threshold as multimodal. We then re-ran the CCA for each threshold to assess stability of results across slightly varying degrees of multimodality.

## Results

### Sample

The final sample of autistic (N=99) and NAI (N=105) did not differ significantly in sex ratio, age, measures of intellectual functioning, measures of structural image quality, number of EEG trials and head motion associated with task- and rs-fMRI (for details see Table 1).

### Unimodal normative models

First, unimodal normative models were estimated. Their accuracy was evaluated using the correlation between the true and the predicted voxel values (Rho), the EV, SMSE and MSLL (Figure S1) and normative models per modality (Figure S3). Evaluation metrics were largely within recommended ranges^92^ and highly similar when modelling age linearly (Figure S2). When testing for group differences in *unimodal* features, there were no significant differences in extreme Z-deviations between autistic and NAI for any of the eight features (Table S4).

### Linked independent component analysis

Next, the Z-deviations (features) were merged using LICA. Fifty independent components were derived across eight different brain feature maps per hemisphere (i.e., modalities) (Figure S5). Overall, across these, the right hemisphere (51.7%) and the left hemisphere (48.3%) did not contribute differentially (χ*²*=1.2, *p*=0.72). Single modality contributions were as follows: EEG R (35.0%) > EEG L (33.2%) > rs-fMRI R (11.2%) > rs-fMRI L (9.6%) > task-fMRI R (3.5%) > task-fMRI L (3.4%) > structure L (2.1%) > structure R (2.1%). Figure S6 shows the correlations between the 50-dimensional factorizations (y-axis) and alternative 40 (Figure S6a) and 60 (Figure S6b) dimensional factorizations. Most components were recovered with high accuracy independently of the order of the factorization. This is in line with previous reports^51^.

### Group Differences

Next, we compared the subject loadings of all (uni- and multimodal) ICs to test for differences between autistic and NAI. Among these, one multimodal IC (#44) showed a significant group difference with autistic individuals having higher contributions compared to NAI (*t*=3.5, *p*_FDR_=0.026) (Figure 2b). There were no significant group differences in the remaining ICs (see Table S5). The significant multimodal component was not differentially driven by the right (52.8%) or left hemisphere (47.2%) (χ*²*=0.4, *p*=0.51) and was associated with several functional modalities (rs-fMRI R [48.5%], rs-fMRI L [35.0%], EEG L [11.6%], task-fMRI R [3.3%]), and to a smaller extent with GM volume (structure R [1.0%], structure L [0.5%]). Figure 3a depicts the spatial and temporal patterns for each imaging modality within IC44. When characterizing these further in terms of their anatomical and functional overlaps with the HOA- and VIS atlases, in the left hemisphere, autistic individuals showed more functional deviations than expected in rs-fMRI connectivity primarily in retinotopic regions of occipital FFG, while to a smaller extent also in lower-order face-selective regions (IOG) (Figures 3b and 3f). In the right hemisphere, they showed linked increased deviations in rs-fMRI and structure primarily in higher-order face-(mFus, pFus) and bodies-selective (OTS) regions of temporal-occipital and occipital FFG (Figures 3c and 3g). On the other hand, regions in the left hemisphere where autistic individuals showed linked decreased deviations compared to NAI, localized to both higher-order face-selective (mFus, pFus) and retinotopic regions of posterior, temporal-occipital and occipital FFG (Figures 3d and 3h). In the right hemisphere, these were mostly in higher-order face face-selective regions (pFus) across rs-fMRI and task-fMRI and in retinotopic regions across structure in temporal-occipital FFG (Figures 3e and 3i). Furthermore, autistic individuals showed more left EEG source activation than expected around 195-203ms and 417-426ms, whereas less source activation at 444–449ms than expected. For further details see Table S6.

**Figure 3.**
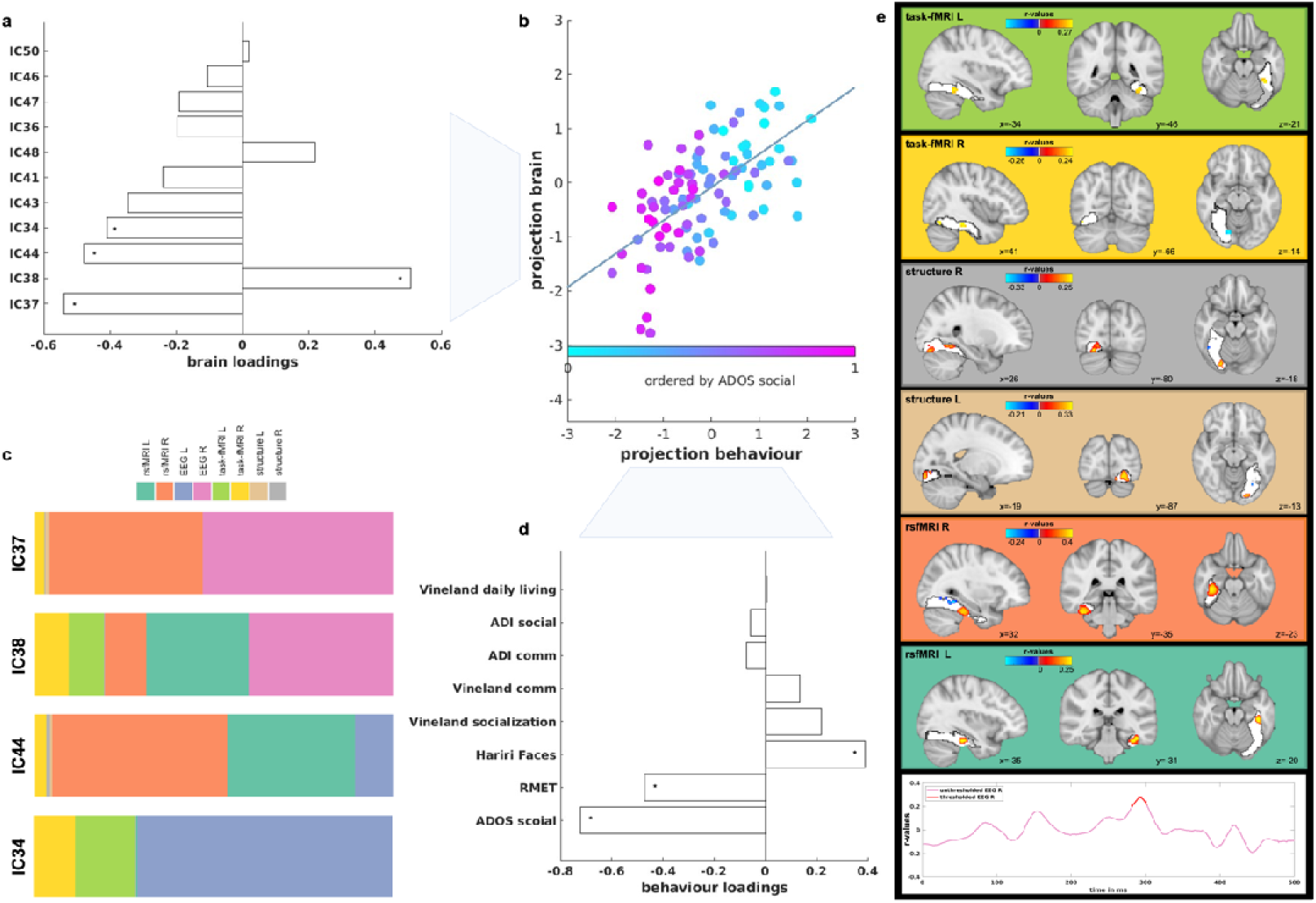
The multivariate association (i.e., canonical correlation) was significant between the eleven multimodal ICs and the social-communicative features associated with autism. Figure 3a shows the loadings of each multimodal component contributing to the CCA mode, while Figure 3d shows the loadings of each social-communicative feature contributing to the CCA mode; stars show the significant loadings. Figure 3b shows the canonical correlation scatterplot color-coded by the highest contributing clinical feature (ADOS social). The x-axis depicts the projected behavioural CCA variate and the y-axis the multimodal ICs CCA variates. Figure 3c shows the modality contributions of the four ICs that contribute significantly to the CCA. Figure 3e depicts the spatial and temporal patterns of each imaging modality that are significantly correlated with the social-communicative features. These are based on the significant correlation values between the Z-deviations of each imaging modality and the canonical imaging variate derived from the CCA.

### Multimodal components

For further analyses, we focused on multimodal components only by excluding those which were primarily driven by one imaging modality, resulting in eleven multimodal ICs (Figure 2a). Across these multimodal ICs, the right hemisphere (60.0%) contributed more than the left hemisphere (40.0%) (χ*²*=7.2, *p*=0.007). Single modality contributions across all multimodal ICs were as follows: EEG R (26.3%) > rs-fMRI L (19.7%) > EEG L (13.9%) > rs-fMRI R (12.1%) > task-fMRI L (9.8%) > task-fMRI R (9.6%) > structure R (4.4%) > structure L (4.2%).

### Autism classification

Applying an SVM, multimodal components performed significantly better at discriminating autistic individuals from NAI (AUC unimodal=0.48, AUC multimodal=0.64, *p*<0.001). This result was confirmed across a range of different multimodality thresholds (see Figure S7a) and was not influenced by varying amounts of features between multimodal and unimodal ICs (Figure S7b).

### Clinical, cognitive associations

The CCA analysis revealed a significant multivariate association between the multimodal ICs and social-communicative features (i.e., ADOS-social affect, ADI-social, ADI-communication, Vineland-Communication, Vineland-Daily Living, Vineland-Socialization, RMET, Hariri face matching condition) (*r*=0.65, *p*_FDR_=0.008; Figure 3b). On the other hand, testing the relationship between the multimodal ICs and non-social features (i.e., ADOS-RRB, ADI-RRB, RBS, SSP, SQ, Hariri shape matching condition), did not yield any significant association (*r*=0.49, *p*_FDR_=0.51; Figure S8) pointing to specificity with social-related features of multimodal ICs. These associations remained stable when varying the multimodality threshold (Figure S9). For the significant association, multimodal IC37 showed the largest contribution on the imaging side followed by IC38, IC44 and IC34 (Figure 3a and 3c), whereas ADOS social-affect, RMET and Hariri face matching scores showed the largest contribution on the behavioural side (Figure 3d). The ICs contributing most are depicted in Figures 2c and Figures S10-12. On average, the right (56.5%) and the left hemisphere (43.5%) did not contribute differentially to these four ICs (χ*²*=2.9, *p*=0.09) which were mostly driven by functional modalities (EEG, task- and rs-fMRI). Next, imaging patterns correlating with social-communication features were characterized in terms of their overlap with anatomical and functional overlaps with the HOA and VIS atlases (Figures 3e and S13). Especially in higher-order face-selective regions (mFus and pFus) of posterior and temporal-occipital FFG, there were both linked *increased* deviations in bilateral rs-fMRI and task-fMRI and linked *decreased* deviations in bilateral structure and right rs-fMRI connectivity. At the same time, particularly in retinotopic regions of occipital FFG there was more bilateral GM volume along with less right task-activation than expected. There were more deviations in right EEG source activation at around 290ms, while left EEG did not reach significance. These joint imaging patterns were associated with more social difficulties as assessed by the ADOS, ADI and Vineland and more errors on the RMET, while also with greater accuracy on the Hariri faces task. For more details, see Tables S6-10.

## Discussion

In the present study, we aimed to characterize the multimodal neural signature of face processing in autism within the FFG, the core region of the face processing network. We identified several ICs that were differentially associated with the four modalities (structure, rs-fMRI, task-fMRI, and EEG), hemispheres, and functional subdivisions of the FFG. Autism-associated differences in FFG organization were more pronounced when penetrating across multiple than single modalities. Furthermore, a set of multimodal ICs was associated with core features related to social functioning, but not non-social functioning, in autism. Taken together, these findings highlight the value of cross-modal analyses in characterizing a key structure in the multilevel neurobiology of autism and its implication in core cognitive and clinical features associated with social functioning.

### Group differences

Among all components, one multimodal component (i.e., IC44) showed a significant difference in subject loadings between autistic and NAI. Overall, the right and left hemisphere did not show differential contributions within this IC, and it was associated with all modalities fed into the analysis, with the functional modalities, especially rs-fMRI and EEG, contributing most (see Figure 2). Particularly, the overlap with the VIS-atlas highlighted that face-selective and retinotopic regions of the FFG were most different between autistic and NAI, again showing a differential pattern by modality and hemisphere. More specifically, in the right hemisphere, higher-order face-selective regions exhibited less task activation and FFA-connectivity than expected, primarily in occipital FFG areas (Figures 2g and 2l). At the same time autistic individuals showed increased deviations in FFA-connectivity primarily in temporal-occipital FFG along with increased GM volume deviations in higher-order face-selective FFG regions (Figures 2e and 2j). This strong right-hemisphere involvement of regions associated with FFA across several modalities is in line with reports of increased FFA volume^38^ and decreased FFA task-activation^93,94^ and FFA-connectivity^34,95^ in autism. Similarly, temporally, autistic individuals also showed more positive left deviations around 195ms which can be indicative of the consistently reported finding of a slower N170 in autistic individuals^36^. This has specifically also been shown and extensively characterised in the same sample^37^. Together these patterns converge to point towards autism-associated differences in face-selective areas of the FFG, both at the structural, functional, and temporal levels. Although these results align with earlier unimodal discoveries, previously it was uncertain whether disparate signals would be separate or coalesce to a joint multimodal expression. In this context, we provide evidence supporting the interconnected nature of distinct signals within a single, unified framework.

On the other hand, in the left hemisphere, IC44-related increased deviations in EEG source activation at around 420ms may indicate reductions in the face-N400 which has been associated with familiar face recognition and semantic information^96,97^. While in NAI face processing becomes the most highly developed visual skill, in autistic individuals faces may convey greater novelty and thus decreased familiarity. Furthermore, in the left hemisphere of IC44, occipital, retinotopic areas of the FFG were most implicated as shown by increased functional connectivity deviations between the FFA and retinotopic and lower-order face-selective areas of the FFG in autistic individuals (Figures 2d and 2i). This was echoed by less GM volume than expected in left hemisphere retinotopic areas of FFG in autistic individuals (Figures 2f and 2k). Retinotopic, early visual areas act as the first stage in a hierarchical network of face processing in which lower-level feature-based components are processed before more complex features in higher-order face-selective regions (e.g., pFus, mFus)^98^. Neural deviations in early visual areas as seen here are in line with reports of autistic individuals showing differences in sensory processing at early perceptual stages and have been described at the cognitive level as weak central coherence^99^. Accordingly, studies show that autistic individuals exhibit a different strategy in processing facial and visual stimuli with a stronger focus on featural, local aspects at the expense of holistic, global information^100^. Similarly, fMRI studies converge to show greater feature-based perceptual strategies in autistic individuals who primarily tend to recruit object-related regions (such as inferior temporal gyrus^101^ or occipital cortex^32^) when viewing facial stimuli. Taken together, this suggests that differences we discovered in the left-hemisphere point primarily to low-level, bottom-up processing differences, whereas in the right-hemisphere they may indicate higher-level atypicalities in the FFA, with a differential involvement across the different structural and functional modalities.

### Clinical, cognitive associations

Multimodal ICs showed a significant multivariate association with a set of clinical and cognitive features associated with social functioning in autism (see Figure 3).

IC44 which also showed a significant group difference was among these significantly contributing ICs to this associations. Components mostly driven by functional modalities (i.e., EEG, rsfMRI and task-fMRI) loaded significantly onto the CCA. Right EEG source activation deviations were at around 280-300ms, potentially indicative of the N250r generated in the FFG^102^ and associated with repetition of familiar facial stimuli^103^. The amplitude of the N250r has been shown to decrease with increasing working memory (WM) load^104^. This would translate into increased deviations as seen in autistic individuals here and may imply differences in degrees of WM resources allocated to the processing of facial stimuli which in turn have a larger novelty character in autistic individuals requiring more attentional effort. With regards to the other modalities, increased deviations particularly in higher-order face-selective regions across brain function (task-fMRI and rs-fMRI) while also in lower-order early visual regions across brain structure were associated with more autistic features, such as more social difficulties as assessed by ADOS, and lower social sensitivity as assessed by the RMET. Previous unimodal studies for example showed that the delayed latency of the N170 predicts change in social adaptive behaviour in autistic individuals^37^ (i.e., EEG), autistic individuals with low performance on facial emotion recognition have reduced bilateral FFG activation (i.e., task-fMRI)^5^ and atypical FFA-connectivity is associated with increased social symptom severity in autism (i.e., rs-fMRI)^34^. Here, we extend unimodal results to a multivariate association across a range of social-communicative features that are related to cross-modal signatures within the FFG. Previously, it was uncertain whether these separate neural signals contribute orthogonally or jointly to social-communicative features in autism. Here, we provide evidence for an interrelated biological basis of core social functioning in autism and that appropriately modelled shared variance across different modalities increases sensitivity to clinical-cognitive features associated with autism^47^. Remarkably, at the same time, there was no association with a set of non-social features, such as repetitive behaviours or sensory processing, pointing to specificity of these multimodal ICs with regards to social functioning.

### Summary and implications

Taken together, the multimodal neural signature within the FFG in autism presents with differential effects across hemispheres, modalities, and topographic organization.

Specifically, the picture emerges that (i) the functional modalities contribute more than the structural modalities and (ii) retinotopic, posterior-occipital regions are more implicated in the left hemisphere and higher-order regions more implicated in the right hemisphere within the FFG when it comes to group differences; but they do not contribute differentially with regards to social functioning. (i) Concurrent neural activity and functional co-expressions (task-, rs-fMRI, EEG) were strongly tied to social features observed in autistic individuals at present (such as current performance and ADOS assessment). On the other hand, more stable structural aspects of the brain established over time and historical symptoms reported through the ADI and Vineland – which provide insights into past behaviors – had a comparatively smaller impact on the observed association. These results highlight the dynamic nature of the relationship between neural activity and social functioning in autism and underscore the importance of considering the temporal dimension when investigating the neural correlates of social functioning in autism. Putative future neuroscientifically informed interventions targeting social features may thus benefit from a focus on concurrent neural functioning. (ii) Topographically, the FFG is known to exhibit an anterior to posterior gradient with more posterior regions related to lower-order, early visual processing, and more anterior regions related to higher-order processing^105^. Here, we see the involvement of both retinotopic and higher-order cognitive, particularly face-sensitive patches pointing to differences in both bottom-up perceptual processes and top-down cognitive information processing in face processing in autism which can amount to a difference in the face processing strategy employed (e.g., more feature-based). These different processing levels are not differentially implicated across hemispheres in the processing of social information in autism suggesting that the distinctive face processing strategy in autism transcends right hemisphere dominance of face processing. On the other hand, hemispheric differences are more apparent in the group-differential IC. Teasing apart hemispheric contributions is particularly important in the light of reports of atypical patterns of brain asymmetry in autistic individuals^30,44,106^. More extreme deviations from a normative model have for example been reported in right temporal-occipital fusiform cortex asymmetry in autistic females^30^, along with more left-lateralized volume in posterior temporal FFG in autistic individuals^40,44^. Subsequent research should delve further into these more nuanced insights revealed by cross-modal analyses pointing to left-lateralized low-level and right-lateralized high-level differences between autistic and NAI.

### Strengths and limitations

Integrating data from different modalities has the advantage of being biologically more informative and comprehensive in characterizing a complex, heterogenous condition like autism. Accordingly, when comparing unimodal deviations in each imaging modality, as well as comparing predominantly unimodal ICs between autistic and NAI, there were no significant group differences, despite employing a more sensitive individual-level measures derived from normative modelling. Also, multimodal features significantly outperformed unimodal features in differentiating autistic form NAI. These results together confirm our hypothesis and previous reports^47,48^ that appropriately modelling cross-modal variance increases sensitivity to detecting subtle effects that may otherwise be missed. Thus, integrating different structural and functional brain measures is the most promising and powerful method to achieve significant advances in our understanding of system-level atypicalities in autism and provides the basis for elucidating mechanisms through which interventions can most efficiently improve clinically relevant functioning^47^. Furthermore, we combine different innovative methods. LICA is particularly powerful when modelling modalities that are different in their numbers of features, spatial correlations, intensity distributions and units. This is, because LICA optimally weighs the contributions of each modality by the correction for the number of effective degrees of freedom and the use of automatic relevance determination priors on components^8,47,49^. Also, by combining normative modelling with LICA, we employ a previously validated approach that has been shown to increase sensitivity in detecting cross-modal effects in clinical populations^52^. Future studies should compare these methods with alternative emerging approaches that aim to combine normative modelling with multimodal integration in a single step, such as the use of variational autoencoders for example^107^.

At the same time, it needs to be pointed out that face processing involves an extended neural network across the whole brain including other structures such as the amygdala, superior temporal sulcus and occipital and frontal cortex^32,52,108,109^. It may thus seem too simplistic to reduce face processing to a single brain region. Still, the FFG has been claimed the core node of a distributed face processing network, as also substantiated by FFG lesion studies^27,110^, and its fine-grained functional heterogeneity warrants careful examination in isolation. Also, implementing cross-modal analyses presents with additional challenges, such as obtaining sufficiently large sample sizes with all participants having available data across all imaging modalities. Here, from a sample of over 600 individuals in the EU-AIMS LEAP dataset, we were able to conduct analysis in just over 200 individuals who had available imaging data across the four different modalities. Whole brain analyses based on multivariate techniques will ultimately require larger sample sizes.

## Conclusion

Integrating information from multiple imaging modalities allows us to gain a more holistic and robust understanding of the complex neural processes underlying core clinical and cognitive features associated with autism. Present results suggest that the FFG is a central region differentially implicated across different neural signals and category-selective regions in autistic and NAI and that this informs cross-modally the mechanisms associated with core social functioning in autism. Eventually, elucidating more precise, integrated and individual-level neural associations of core cognitive and clinical features, will pave the way for further work identifying stratification, mechanistic and prognostic biomarkers, and the development of more personalised support, thereby eventually improving the quality of lives of autistic individuals.

## Data Availability

Data produced in the present study are available upon reasonable request to the authors.
Code is available at: https://github.com/allera/Llera_elife_2019_1/tree/master/matlab_flica_toolbox and https://github.com/amarquand/PCNtoolkit.

## Acknowledgements

We thank all participants and their families for participating in the studies that contribute to the datasets used in this research. We also gratefully acknowledge the contributions of all members of the EU-AIMS/AIMS-2-TRIALS LEAP group: Jumana Ahmad, Sara Ambrosino, Bonnie Auyeung, Sarah Baumeister, Sven Bölte, Carsten Bours, Michael Brammer, Daniel Brandeis, Claudia Brogna, Yvette de Bruijn, Bhismadev Chakrabarti, Ineke Cornelissen, Daisy Crawley, Guillaume Dumas, Jessica Faulkner, Vincent Frouin, Pilar Garcés, David Goyard, Lindsay Ham, Hannah Hayward, Joerg Hipp, Mark H. Johnson, Emily J.H. Jones, Xavier Liogier D’ardhuy, David J. Lythgoe, René Mandl, Luke Mason, Andreas Meyer-Lindenberg, Nico Mueller, Bethany Oakley, Laurence O’Dwyer, Bob Oranje, Gahan Pandina, Antonio M. Persico, Barbara Ruggeri, Amber Ruigrok, Jessica Sabet, Roberto Sacco, Antonia San José Cáceres, Emily Simonoff, Will Spooren, Roberto Toro, Heike Tost, Jack Waldman, Steve C.R. Williams, Caroline Wooldridge, and Marcel P. Zwiers. This project has received funding from the Innovative Medicines Initiative 2 Joint Undertaking under grant agreement No 115300 (for EU-AIMS) and No 777394 (for AIMS-2-TRIALS). This Joint Undertaking receives support from the European Union’s Horizon 2020 research and innovation programme and EFPIA and AUTISM SPEAKS, Autistica, SFARI. Any views expressed are those of the author(s) and not necessarily those of the funders (IHI-JU2). This work was also supported by the Netherlands Organization for Scientific Research through Vidi grants (Grant No. 864.12.003 [to CFB]; from the FP7 (Grant Nos. 602805) (AGGRESSOTYPE) (to JKB), 603016 (MATRICS), and 278948 (TACTICS); and from the European Community’s Horizon 2020 Programme (H2020/2014-2020) (Grant Nos. 643051 [MiND] and 642996 (BRAINVIEW). This work received funding from the Wellcome Trust UK Strategic Award (Award No. 098369/Z/12/Z) and from the National Institute for Health Research Maudsley Biomedical Research Centre (to DM). DLF is supported by funding from the European Union’s Horizon 2020 research and innovation programme under the Marie Skłodowska-Curie grant agreement No 101025785. EJHJ and RH received funding from SFARI GAIINS (grant number 10039678). SB-C is funded by the Autism Research Trust, the Wellcome Trust, the Templeton World Charitable Foundation and by the NIHR Biomedical Research Centre in Cambridge, during the period of this work. BHV is supported by the Swiss National Science Foundation [10001C_197480].

## References

1. Maenner, M. J. et al. Prevalence and Characteristics of Autism Spectrum Disorder Among Children Aged 8 Years - Autism and Developmental Disabilities Monitoring Network, 11 Sites, United States, 2018. MMWR Surveill Summ 70, 1–16 (2021).

2. Kanne, S. M. et al. The role of adaptive behavior in autism spectrum disorders: Implications for functional outcome. Journal of Autism and Developmental Disorders 41, (2011).

3. Dawson, G., Webb, S. J. & McPartland, J. Understanding the nature of face processing impairment in autism: insights from behavioral and electrophysiological studies. Dev Neuropsychol 27, 403–424 (2005).

4. Sasson, N. J. The development of face processing in autism. J Autism Dev Disord 36, 381–394 (2006).

5. Meyer-Lindenberg, H. et al. Facial expression recognition is linked to clinical and neurofunctional differences in autism. Molecular Autism 13, 43 (2022).

6. Chevallier, C., Huguet, P., Happé, F., George, N. & Conty, L. Salient Social Cues are Prioritized in Autism Spectrum Disorders Despite Overall Decrease in Social Attention. J Autism Dev Disord 43, 1642–1651 (2013).

7. Schultz, R. T. Developmental deficits in social perception in autism: the role of the amygdala and fusiform face area. International Journal of Developmental Neuroscience 23, 125–141 (2005).

8. Sui, J., Adali, T., Yu, Q., Chen, J. & Calhoun, V. D. A review of multivariate methods for multimodal fusion of brain imaging data. Journal of Neuroscience Methods 204, 68– 81 (2012).

9. Grill-Spector, K. & Weiner, K. S. The functional architecture of the ventral temporal cortex and its role in categorization. Nat Rev Neurosci 15, 536–548 (2014).

10. Kanwisher, N., Stanley, D. & Harris, A. The fusiform face area is selective for faces not animals. NeuroReport 10, 183 (1999).

11. Kanwisher, N., McDermott, J. & Chun, M. M. The Fusiform Face Area: A Module in Human Extrastriate Cortex Specialized for Face Perception. J. Neurosci. 17, 4302–4311 (1997).

12. Clark, V. P. et al. Functional Magnetic Resonance Imaging of Human Visual Cortex during Face Matching: A Comparison with Positron Emission Tomography. NeuroImage 4, 1–15 (1996).

13. McCarthy, G., Puce, A., Gore, J. C. & Allison, T. Face-Specific Processing in the Human Fusiform Gyrus. Journal of Cognitive Neuroscience 9, 605–610 (1997).

14. Kanwisher, N. & Yovel, G. The fusiform face area: a cortical region specialized for the perception of faces. Philosophical Transactions of the Royal Society B: Biological Sciences 361, 2109–2128 (2006).

15. Bentin, S., Allison, T., Puce, A., Perez, E. & McCarthy, G. Electrophysiological Studies of Face Perception in Humans. Journal of Cognitive Neuroscience 8, 551–565 (1996).

16. Bötzel, K., Schulze, S. & Stodieck, S. R. Scalp topography and analysis of intracranial sources of face-evoked potentials. Exp Brain Res 104, 135–143 (1995).

17. Engell, A. D. & McCarthy, G. Face, eye, and body selective responses in fusiform gyrus and adjacent cortex: an intracranial EEG study. Frontiers in Human Neuroscience 8, (2014).

18. Rosenke, M., van Hoof, R., van den Hurk, J., Grill-Spector, K. & Goebel, R. A Probabilistic Functional Atlas of Human Occipito-Temporal Visual Cortex. Cerebral Cortex 31, 603–619 (2021).

19. Zhang, W. et al. Functional organization of the fusiform gyrus revealed with connectivity profiles. Hum Brain Mapp 37, 3003–3016 (2016).

20. Gauthier, I. et al. The fusiform ‘face area’ is part of a network that processes faces at the individual level. Journal of Cognitive Neuroscience (2000) doi:10.1162/089892900562165.

21. Taylor, J. C., Wiggett, A. J. & Downing, P. E. Functional MRI Analysis of Body and Body Part Representations in the Extrastriate and Fusiform Body Areas. Journal of Neurophysiology 98, 1626–1633 (2007).

22. Wang, X. et al. Where color rests: Spontaneous brain activity of bilateral fusiform and lingual regions predicts object color knowledge performance. NeuroImage 76, 252–263 (2013).

23. Mion, M. et al. What the left and right anterior fusiform gyri tell us about semantic memory. Brain 133, 3256–3268 (2010).

24. Balsamo, L. M., Xu, B. & Gaillard, W. D. Language lateralization and the role of the fusiform gyrus in semantic processing in young children. NeuroImage 31, 1306–1314 (2006).

25. Rossion, B. et al. Hemispheric Asymmetries for Whole-Based and Part-Based Face Processing in the Human Fusiform Gyrus. Journal of Cognitive Neuroscience 12, 793– 802 (2000).

26. Yovel, G., Tambini, A. & Brandman, T. The asymmetry of the fusiform face area is a stable individual characteristic that underlies the left-visual-field superiority for faces. Neuropsychologia 46, 3061–3068 (2008).

27. Rossion, B. et al. A network of occipito-temporal face-sensitive areas besides the right middle fusiform gyrus is necessary for normal face processing. Brain (2003) doi:10.1093/brain/awg241.

28. Niina, M., Okamura, J. & Wang, G. Electrophysiological evidence for separation between human face and non-face object processing only in the right hemisphere. International Journal of Psychophysiology 98, 119–127 (2015).

29. Cardinale, R. C., Shih, P., Fishman, I., Ford, L. M. & Müller, R. A. Pervasive rightward asymmetry shifts of functional networks in autism spectrum disorder. JAMA Psychiatry (2013) doi:10.1001/jamapsychiatry.2013.382.

30. Floris, D. L. et al. Atypical Brain Asymmetry in Autism—A Candidate for Clinically Meaningful Stratification. Biological Psychiatry: Cognitive Neuroscience and Neuroimaging 6, 802–812 (2021).

31. Nickl-Jockschat, T. et al. Neural networks related to dysfunctional face processing in autism spectrum disorder. Brain Struct Funct 220, 2355–2371 (2015).

32. Pierce, K., Müller, R. A., Ambrose, J., Allen, G. & Courchesne, E. Face processing occurs outside the fusiform ‘face area’ in autism: Evidence from functional MRI. Brain 124, (2001).

33. Pierce, K., Haist, F., Sedaghat, F. & Courchesne, E. The brain response to personally familiar faces in autism: findings of fusiform activity and beyond. Brain 127, 2703–2716 (2004).

34. Kleinhans, N. M. et al. Abnormal functional connectivity in autism spectrum disorders during face processing. Brain 131, 1000–1012 (2008).

35. Koshino, H. et al. fMRI Investigation of Working Memory for Faces in Autism: Visual Coding and Underconnectivity with Frontal Areas. Cerebral Cortex 18, 289–300 (2008).

36. Kang, E. et al. Atypicality of the N170 Event-Related Potential in Autism Spectrum Disorder: A Meta-analysis. Biological Psychiatry: Cognitive Neuroscience and Neuroimaging 3, 657–666 (2018).

37. Mason, L. et al. Stratifying the autistic phenotype using electrophysiological indices of social perception. Science Translational Medicine 14, eabf8987 (2022).

38. Waiter, G. D. et al. A voxel-based investigation of brain structure in male adolescents with autistic spectrum disorder. NeuroImage 22, (2004).

39. Van Kooten, I. A. J. et al. Neurons in the fusiform gyrus are fewer and smaller in autism. Brain 131, (2008).

40. Herbert, M. R. et al. Abnormal asymmetry in language association cortex in autism. Annals of neurology 52, 588–96 (2002).

41. Neuhaus, E. et al. The Relationship Between Early Neural Responses to Emotional Faces at Age 3 and Later Autism and Anxiety Symptoms in Adolescents with Autism. J Autism Dev Disord 46, 2450–2463 (2016).

42. Webb, S. J. et al. Developmental change in the ERP responses to familiar faces in toddlers with autism spectrum disorders versus typical development. Child Dev 82, 1868–1886 (2011).

43. Gao, C., Conte, S., Richards, J. E., Xie, W. & Hanayik, T. The neural sources of N170: Understanding timing of activation in face-selective areas. Psychophysiology 56, e13336 (2019).

44. Dougherty, C. C., Evans, D. W., Katuwal, G. J. & Michael, A. M. Asymmetry of fusiform structure in autism spectrum disorder: Trajectory and association with symptom severity. Molecular Autism 7, (2016).

45. Iidaka, T., Matsumoto, A., Haneda, K., Okada, T. & Sadato, N. Hemodynamic and electrophysiological relationship involved in human face processing: Evidence from a combined fMRI–ERP study. Brain and Cognition 60, 176–186 (2006).

46. Puce, A. et al. The human temporal lobe integrates facial form and motion: evidence from fMRI and ERP studies. NeuroImage 19, 861–869 (2003).

47. Groves, A. R. et al. Benefits of multi-modal fusion analysis on a large-scale dataset: Life-span patterns of inter-subject variability in cortical morphometry and white matter microstructure. NeuroImage 63, (2012).

48. Calhoun, V. D. & Sui, J. Multimodal Fusion of Brain Imaging Data: A Key to Finding the Missing Link(s) in Complex Mental Illness. Biological Psychiatry: Cognitive Neuroscience and Neuroimaging 1, 230–244 (2016).

49. Groves, A. R., Beckmann, C. F., Smith, S. M. & Woolrich, M. W. Linked independent component analysis for multimodal data fusion. NeuroImage 54, 2198–2217 (2011).

50. Francx, W. et al. Integrated analysis of gray and white matter alterations in attention-deficit/hyperactivity disorder. NeuroImage: Clinical 11, 357–367 (2016).

51. Llera, A., Wolfers, T., Mulders, P. & Beckmann, C. F. Inter-individual differences in human brain structure and morphology link to variation in demographics and behavior. eLife 8, (2019).

52. Holz, N. E. et al. Age-related brain deviations and aggression. Psychol Med 1–10 (2022) doi:10.1017/S003329172200068X.

53. Brown, T. T. et al. Neuroanatomical assessment of biological maturity. Current Biology 22, (2012).

54. Erus, G. et al. Imaging patterns of brain development and their relationship to cognition. Cerebral Cortex 25, (2015).

55. Liem, F. et al. Predicting brain-age from multimodal imaging data captures cognitive impairment. NeuroImage 148, (2017).

56. Mei, T. et al. Autism Is Associated With Interindividual Variations of Gray and White Matter Morphology. Biol Psychiatry Cogn Neurosci Neuroimaging S2451-9022(22)00212–9 (2022) doi:10.1016/j.bpsc.2022.08.011.

57. Oblong, L. M. et al. Linking functional and structural brain organisation with behaviour in autism: a multimodal EU-AIMS Longitudinal European Autism Project (LEAP) study. Molecular Autism 14, 32 (2023).

58. Charman, T. et al. The EU-AIMS Longitudinal European Autism Project (LEAP): Clinical characterisation. Molecular Autism 8, (2017).

59. Loth, E. et al. The EU-AIMS Longitudinal European Autism Project (LEAP): Design and methodologies to identify and validate stratification biomarkers for autism spectrum disorders. Molecular Autism 8, (2017).

60. Marquand, A. F., Rezek, I., Buitelaar, J. & Beckmann, C. F. Understanding Heterogeneity in Clinical Cohorts Using Normative Models: Beyond Case-Control Studies. Biological Psychiatry 80, 552–561 (2016).

61. Parkes, L. et al. Transdiagnostic dimensions of psychopathology explain individuals’ unique deviations from normative neurodevelopment in brain structure. Transl Psychiatry 11, 232 (2021).

62. Lord, C., Rutter, M. & Le Couteur, A. Autism Diagnostic Interview-Revised: a revised version of a diagnostic interview for caregivers of individuals with possible pervasive developmental disorders. Journal of autism and developmental disorders 24, 659–685 (1994).

63. Lord, C. et al. The autism diagnostic observation schedule-generic: a standard measure of social and communication deficits associated with the spectrum of autism. Journal of autism and developmental disorders 30, 205–223 (2000).

64. Sparrow, S. S., Cicchetti, D. V. & Balla, D. A. The Vineland Adaptive Behavior Scales (2nd ed). in Major psychological assessment instruments (2005).

65. Hariri, A. R., Tessitore, A., Mattay, V. S., Fera, F. & Weinberger, D. R. The amygdala response to emotional stimuli: A comparison of faces and scenes. NeuroImage 17, (2002).

66. Baron-Cohen, S., Wheelwright, S., Hill, J., Raste, Y. & Plumb, I. The ‘Reading the Mind in the Eyes’ Test revised version: A study with normal adults, and adults with Asperger syndrome or high-functioning autism. Journal of Child Psychology and Psychiatry and Allied Disciplines 42, (2001).

67. Bodfish, J. W., Symons, F. J., Parker, D. E. & Lewis, M. H. Repetitive Behavior Scale– Revised. PsycTESTS® (2000).

68. Auyeung, B. et al. The children’s empathy quotient and systemizing quotient: Sex differences in typical development and in autism spectrum conditions. Journal of Autism and Developmental Disorders 39, (2009).

69. Auyeung, B., Allison, C., Wheelwright, S. & Baron-Cohen, S. Brief report: Development of the adolescent empathy and systemizing quotients. Journal of Autism and Developmental Disorders 42, (2012).

70. Baron-Cohen, S., Richler, J., Bisarya, D., Gurunathan, N. & Wheelwright, S. The systemizing quotient: An investigation of adults with Asperger syndrome or high-functioning autism, and normal sex differences. Philosophical Transactions of the Royal Society B: Biological Sciences 358, (2003).

71. Tomchek, S. D. & Dunn, W. Sensory processing in children with and without autism: a comparative study using the short sensory profile. American Journal of Occupational Therapy 61, (2007).

72. Llera, A. et al. Evaluation of data imputation strategies in complex, deeply-phenotyped data sets: the case of the EU-AIMS Longitudinal European Autism Project. BMC Medical Research Methodology 22, 229 (2022).

73. Looden, T. et al. Patterns of connectome variability in autism across five functional activation tasks: findings from the LEAP project. Molecular Autism 13, 53 (2022).

74. Ashburner, J. A fast diffeomorphic image registration algorithm. NeuroImage 38, 95–113 (2007).

75. Pruim, R. H. R. et al. ICA-AROMA: A robust ICA-based strategy for removing motion artifacts from fMRI data. NeuroImage 112, 267–277 (2015).

76. Parkes, L., Fulcher, B., Yücel, M. & Fornito, A. An evaluation of the efficacy, reliability, and sensitivity of motion correction strategies for resting-state functional MRI. NeuroImage 171, 415–436 (2018).

77. Greve, D. N. & Fischl, B. Accurate and robust brain image alignment using boundary-based registration. NeuroImage 48, 63–72 (2009).

78. Delorme, A. & Makeig, S. EEGLAB: an open source toolbox for analysis of single-trial EEG dynamics including independent component analysis. J Neurosci Methods 134, 9– 21 (2004).

79. Oostenveld, R., Fries, P., Maris, E. & Schoffelen, J.-M. FieldTrip: Open source software for advanced analysis of MEG, EEG, and invasive electrophysiological data. Comput Intell Neurosci 2011, 156869 (2011).

80. Marquand, A. F. et al. Conceptualizing mental disorders as deviations from normative functioning. Molecular Psychiatry 24, 1415–1424 (2019).

81. Rutherford, S. et al. Charting brain growth and aging at high spatial precision. eLife 11, e72904 (2022).

82. Fraza, C. J., Dinga, R., Beckmann, C. F. & Marquand, A. F. Warped Bayesian linear regression for normative modelling of big data. NeuroImage 245, 118715 (2021).

83. Holz, N. E. et al. A stable and replicable neural signature of lifespan adversity in the adult brain. Nat Neurosci 1–10 (2023) doi:10.1038/s41593-023-01410-8.

84. Wolfers, T. et al. Mapping the Heterogeneous Phenotype of Schizophrenia and Bipolar Disorder Using Normative Models. JAMA Psychiatry 75, 1146–1155 (2018).

85. Wolfers, T. et al. Individual differences v. the average patient: Mapping the heterogeneity in ADHD using normative models. Psychological Medicine 50, 314–323 (2019).

86. Wolfers, T. et al. Refinement by integration: Aggregated effects of multimodal imaging markers on adult ADHD. Journal of Psychiatry and Neuroscience 42, 386–394 (2017).

87. Floris, D. L. & Howells, H. Atypical structural and functional motor networks in autism. in Progress in Brain Research: Cerebral lateralization and cognition: Evolutionary and developmental investigations of behavioral biases (eds. Forrester, G. S., Hopkins, W. D., Hudry, K. & Lindell, A. K.) vol. 238 207–248 (Elsevier Academic Press, 2018).

88. Benjamini, Y. & Hochberg, Y. On the adaptive control of the false discovery rate in multiple testing with independent statistics. Journal of Educational and Behavioral Statistics (2000) doi:10.3102/10769986025001060.

89. Hotelling, H. Relations Between Two Sets of Variates. Biometrika 28, 321–377 (1936).

90. Winkler, A. M., Renaud, O., Smith, S. M. & Nichols, T. E. Permutation inference for canonical correlation analysis. NeuroImage 220, (2020).

91. Ball, G. et al. Multimodal image analysis of clinical influences on preterm brain development. Annals of Neurology 82, 233–246 (2017).

92. Rutherford, S. et al. Evidence for embracing normative modeling. eLife 12, e85082 (2023).

93. Critchley, H. D. et al. The functional neuroanatomy of social behaviour: changes in cerebral blood flow when people with autistic disorder process facial expressions. Brain 123 **( Pt** **11**), 2203–2212 (2000).

94. Corbett, B. A. et al. A functional and structural study of emotion and face processing in children with autism. Psychiatry Research: Neuroimaging 173, 196–205 (2009).

95. Lynn, A. C. et al. Functional connectivity differences in autism during face and car recognition: underconnectivity and atypical age-related changes. Developmental Science 21, e12508 (2018).

96. Bentin, S. & Deouell, L. Y. Structural Encoding and Identification in Face Processing: Erp Evidence for Separate Mechanisms. Cognitive Neuropsychology 17, 35–55 (2000).

97. Eimer, M. Event-related brain potentials distinguish processing stages involved in face perception and recognition. Clinical Neurophysiology 111, 694–705 (2000).

98. Pitcher, D., Walsh, V. & Duchaine, B. The role of the occipital face area in the cortical face perception network. Exp Brain Res 209, 481–493 (2011).

99. Happé, F. & Frith, U. The weak coherence account: detail-focused cognitive style in autism spectrum disorders. Journal of autism and developmental disorders 36, 5–25 (2006).

100. Watson, T. L. Implications of holistic face processing in autism and schizophrenia. Front Psychol 4, 414 (2013).

101. Schultz, R. T. et al. Abnormal Ventral Temporal Cortical Activity During Face Discrimination Among Individuals With Autism and Asperger Syndrome. Archives of General Psychiatry 57, 331–340 (2000).

102. Schweinberger, S. R., Pickering, E. C., Jentzsch, I., Burton, A. M. & Kaufmann, J. M. Event-related brain potential evidence for a response of inferior temporal cortex to familiar face repetitions. Cognitive Brain Research 14, 398–409 (2002).

103. Schweinberger, S. R., Huddy, V. & Burton, A. M. N250r: a face-selective brain response to stimulus repetitions. NeuroReport 15, 1501 (2004).

104. Morgan, H. M., Klein, C., Boehm, S. G., Shapiro, K. L. & Linden, D. E. J. Working memory load for faces modulates P300, N170, and N250r. J Cogn Neurosci 20, 989– 1002 (2008).

105. Caspers, J. et al. Functional characterization and differential coactivation patterns of two cytoarchitectonic visual areas on the human posterior fusiform gyrus. Human Brain Mapping 35, 2754–2767 (2014).

106. Floris, D. L. et al. Atypically rightward cerebral asymmetry in male adults with autism stratifies individuals with and without language delay. Human Brain Mapping 37, 230–253 (2016).

107. Lawry Aguila, A., Chapman, J. & Altmann, A. Multi-modal Variational Autoencoders for Normative Modelling Across Multiple Imaging Modalities. in Medical Image Computing and Computer Assisted Intervention – MICCAI 2023 (eds. Greenspan, H. et al.) 425–434 (Springer Nature Switzerland, 2023). doi:10.1007/978-3-031-43907-0_41.

108. Aoki, Y., Cortese, S. & Tansella, M. Neural bases of atypical emotional face processing in autism: A meta-analysis of fMRI studies. World Journal of Biological Psychiatry 16, (2015).

109. Haxby, J. V., Hoffman, E. A. & Gobbini, M. I. The distributed human neural system for face perception. Trends Cogn Sci 4, 223–233 (2000).

110. Barton, J. J. S., Press, D. Z., Keenan, J. P. & O’Connor, M. Lesions of the fusiform face area impair perception of facial configuration in prosopagnosia. Neurology 58, 71– 78 (2002).

