## Supplement for "A multimodal neural signature of face processing in autism within the fusiform gyrus"

**Supplementary Information**

### Supplementary Methods

### Participants, study design and exclusion criteria

Full details of the Longitudinal European Autism Project (LEAP) study design can be found in ^1,2^. The LEAP protocols are available at https://www.eu-aims.eu/fileadmin/websites/eu-aims/media/EU-AIMS_LEAP/EU-AIMS-LEAP_SOP_StudyProtocol.zip. The study was approved by the local ethical committees of participating centres, and written informed consent was obtained from all participants or their legal guardians (for participants <18 years). All autistic participants had an existing clinical diagnosis of autism according to DSM-IV^3^, DSM-IV-TR^4^, DSM-5^5^ or ICD-10^6^ criteria. In the autism group, diagnosis was confirmed using the combined information of gold-standard diagnostic instruments, the Autism Diagnostic Interview-Revised^7^ (ADI-R) and the Autism Diagnostic Observation Schedule (ADOS)^8^. Appropriate to a multi-centre study, quality control procedures were in place around training, and data collection/entry. Cross-site training sessions for collecting clinical data were put in place, the ADOS and ADI-R were administered and scored by qualified/certified personnel and the study was regularly monitored according to Good Clinical Practice (GCP) standards. Participants underwent comprehensive clinical, cognitive and MRI assessment at one of six collaborating sites: the Institute of Psychiatry, Psychology and Neurosciences, King’s College London (KCL), London, United Kingdom; Autism Research Centre at the University of Cambridge, Cambridge, United Kingdom; Radboud University Nijmegen Medical Centre, Nijmegen, the Netherlands; University Medical Centre Utrecht, Utrecht, the Netherlands; Central Institute of Mental Health, Mannheim, Germany; and University Campus Bio-Medico, Rome, Italy. Exclusion criteria included the presence of any MRI contraindications (e.g., metal implants, braces, claustrophobia) or failure to give informed written consent to MRI scanning, as well as significant hearing or visual impairments not corrected by glasses or hearing aids. In addition to general exclusion criteria, we also excluded individuals based on different sets of criteria per each imaging modality as follows:

- **Structural T1-weighted images** with excessive head motion (N=37), clinically (mostly) non-significant atypicalities (N=20) and failed preprocessing (N=2) were excluded. Autistic and non-autistic individuals did not differ on measures of image quality derived from SPM after exclusions (*W*=4926, *p*=0.52).
- **Resting-state fMRI data** (rs-fMRI) was available for five out of the six acquisition sites (not collected for the Rome site). Scans with less than 180 volumes (i.e., less than 90% of R-fMRI scan completed; N=8), a TR different than the standard TR =2.3 (N=2), excessive head motion during the R-fMRI scan (N=29 with mean root-mean-square of the framewise displacement [meanFD] >0.7 and N= 38 with maxFD >7.6 [i.e., motion of more than 2 voxels]), low full-brain coverage (N=24), extreme values (N=12 with values >4SD) and failed preprocessing (N=1) were excluded. Autistic and non-autistic individuals did not differ on meanFD after exclusions (*W*=4915, *p*=0.5).
- **Hariri task-fMRI** datasets with excessive head motion (>20% of trials with a meanFD greater than 0.5 mm; N=47) were excluded. Autistic and non-autistic individuals did not differ on the percentage of trials with a meanFD greater than 0.5mm after exclusions (*W*=5279, *p*=0.83).
- **EEG data** was available for five out of the six acquisition sites (not collected for the Cambridge site). Images with technical/upload errors (N=63), incomplete EEG battery (N=32), too few trials (<20 artifact-free; N=9) and too few channels (<75%; N=2) were excluded.

Applying all exclusion criteria resulted in a sample of 204 individuals (99 autism, 105 non-autistic individuals [NAI]) between 7 and 30 years of age matched for age, full-scale IQ (FIQ), verbal IQ (VIQ), performance IQ (PIQ), structural image quality and head motion in rs-fMRI and task-fMRI across diagnostic groups. For details see Table 1.

### Demographic clinical, and cognitive measures

#### Intellectual functioning

General intellectual abilities were assessed using the Wechsler Abbreviated Scales of Intelligence-Second Edition^9^ (WASI-II), or if unavailable the Wechsler Intelligence Scale for Children-III/IV^10,11^ (WISC-III/IV) for children or Wechsler Adult Intelligence Scale for Adults-III/IV^12,13^ (WAIS-III/IV) for adults. Standardized estimates of VIQ, PIQ, and FIQ were derived using IQ norms with mean=100 and SD=±15.

#### ADOS

The Autism Diagnostic Observation Schedule^8^ (ADOS-G) was used to measure the impact of current, clinically observed core symptoms of autism. Based on ADOS-2 algorithm totals^14,15^, we report ADOS-2 Calibrated Severity Score (CSS) for ‘Social Affect’ indexing social-communication difficulties and ‘RRBs’ indexing restricted and repetitive behaviours. The CSS scores range from 1 to 10, with higher scores indicating more severe ASD symptom severity.

#### ADI-R

The Autism Diagnostic Interview-Revised^7^ (ADI-R) is a structured parent interview completed by parents or caregivers of participants with autism. Algorithm scores were derived from current and historical symptom information for the domains of Reciprocal Social Interaction, Communication, and Restricted, Repetitive and Stereotyped Behaviours and Interests.

#### Vineland

Adaptive behaviour was assessed with semi-structured parent/carer interviews using the Vineland Adaptive Behavior Scale-Second Edition. This measures a person’s current level of functioning across three domains: communication (expressive, receptive, and written), daily living skills (community, domestic, and personal), and socialization (coping skills, interpersonal relationships, and play and leisure time). For each domain, standard scores were obtained and combined to generate an Adaptive Behaviour Composite (ABC) score. Standard scores have a mean of 100 (SD = 15), with lower scores indicating greater functional impairment.

#### Emotional face matching task (Hariri)

Participants completed a well-established face matching task^16^ within the MRI scanner, with alternating blocks of faces (showing angry and fearful emotions) and control conditions. In the emotional face condition, a target face has to be matched to one of two probes (identity match) by pressing the left or right button of a response device. Analogously, in the control condition, participants are asked to match a target shape (circle or ellipses) to two test shapes. Behavioural performance on the task was extracted as the accuracy on performing the task (i.e., percentage of successful trials) and used as our and used as independent variable for our analyses.

#### The Reading the Mind in the Eyes Test

The Reading the Mind in the Eyes test^17^ asks participants to identify complex emotions and mental states based only on the eye region of a face. Depending on their age (adults 18-30, adolescents 12-17, children 6-11) and ability level, participants received either an adult (36 items), adolescent (31 items) or child (28 items) version of the test. Percentage of correct answers was used as the outcome variable. Before merging the three different versions across all subjects, each age-related version was z-standardized.

#### Systemizing Quotient

The Systemizing Quotient (SQ) was used to measure a cognitive style characterised by the motivation to predict lawful events (using if-then rules) and observations of input-operation-output relationships and includes good attention to detail. Age-appropriate versions were used for children^18^, adolescents^19^ and adults^20^. Before merging the three different versions across all individuals, each version was z-standardized.

#### Repetitive Behaviour Scale

The Repetitive Behaviour Scale‐Revised^21^ (RBS‐R), composed of 43 items, was used to derive parent‐reported total raw scores for restricted and repetitive behaviours, with higher scores indicating a greater level of atypical behaviours.

#### Short Sensory Profile

Sensory processing atypicalities were assessed using the Short Sensory Profile^22^ (SSP) across 38 items, from which a total raw score was obtained (lower scores indicate more atypicality) that reflect dysfunction across multiple sensory domains.

While ADOS and ADI were only present in autistic individuals, the remaining measures were collected in both autistic and non-autistic individuals. The canonical correlation analysis examining the relationship between multimodal components and clinical cognitive measures was conducted in autistic individuals only.

To test the specificity of face processing related independent components with measures related to social-communication specifically, but not with non-social features, we split available autism-associated measures into two sets of feature sets: 1) Social-communicative features comprising measures of difficulties with social communication and daily living skills (i.e., ADOS-SA, ADI-communication, ADI-social, Vineland scales), emotional face matching performance (i.e., Hariri faces task), and social sensitivity to complex emotions (i.e., RMET) and 2) non-social features comprising restricted, repetitive behaviours (i.e., ADOS-RRB, ADI-RRB, RBS-R), systemizing (i.e., SQ), shape matching performance (i.e., Hariri shapes task, as the control condition to the Hariri emotional faces task) and sensory processing atypicalities (i.e., SSP). See Table S1 for further details.

To tackle missing clinical data and to not further reduce sample size, we used imputed clinical data^23^, as in previous work with this dataset^24,25^. The imputation procedure considered the potential non-randomness of missing data, and therefore developed quantitative measures to assess the quality of the imputations, and finally imputed data adopting a nonparametric tree regression model embedded in an iterative round-robin iterative schedule. The exact procedure within this dataset is published^23^.

### MRI and EEG data acquisition

MRI data were acquired on 3T scanners: General Electric MR750 (GE Medical Systems, Milwaukee, WI, USA) at Institute of Psychiatry, Psychology and Neuroscience, King’s College London, United Kingdom (KCL); Siemens Magnetom Skyra (Siemens, Erlangen, Germany) at Radboud University Nijmegen Medical Centre, the Netherlands (RUNMC); Siemens Magnetom Verio (Siemens, Erlangen, Germany) at Autism Research Centre at the University of Cambridge, United Kingdom (UCAM); Philips 3T Achieva (Philips Healthcare Systems, Best, The Netherlands) at University Medical Centre Utrecht, the Netherlands (UMCU); GE Medical Systems Signa HDxTt at the Rome University; and Siemens Magnetom Trio (Siemens, Erlangen, Germany) at Central Institute of Mental Health, Mannheim, Germany (CIMH). Procedures were undertaken to optimize the MRI sequences for the best scanner-specific options, and phantoms and travelling heads were employed to assure standardization and quality assurance of the multi-site image-acquisition.

**T1-weighted images**: images were obtained using a 5.5 minute MPRAGE sequence (Nijmegen site example: TR=2300ms, TE=2.93ms, T1=900ms, voxels size=1.1x1.1x1.2mm, flip angle=9°, matrix size=256x256, FOV=270mm, 176slices). Slight variations are present across centres, for details see Table S1.

**Resting-state fMRI**: An eight-to- ten minute rs-fMRI scan was acquired using a multi-echo planar imaging (ME-EPI) sequence developed by Kundu et al.^26^; TR=2300ms, TE~12ms, 31ms, and 48ms (slight variations are present across centres), flip angle=80°, matrix size=64x64, in-plane resolution=3.8mm, FOV=240mm, 33 axial slices, slice thickness/gap=3.8mm/0.4mm, volumes=200 (UMCU), 215 (KCL, CIMH), or 265 (RUNMC, UCAM). Participants were instructed to relax and fixate on a cross presented on the screen for the duration of the R-fMRI scan. For further details, see Table S2.

**Task-fMRI**: Data were acquired with echo-planar T2*-weighted imaging (EPI), sensitive to the Blood Oxygenation Level Dependent (BOLD) signal contrast. We used a well-established task that probe functional brain responses during socio-emotional information processing. In the face-matching task, participants were asked to perform an identity match of a target stimulus to one of two probes presented underneath the target. In the faces condition, stimuli consisted of fearful or angry faces. In the control condition, stimuli consisted of geometric shapes. Responses were recorded as button press. The task was presented in eight blocks of six trials of 30s each, with alternating blocks of faces and shapes (total duration: 4:28 min).

**EEG**: five sites acquired EEG data in LEAP: Kings College, London (KCL), The Central Institute of Mental Health, Mannheim (CIMH), University Medical Centre, Utrecht (UMCU), Radboud University Nijmegen Medical Centre (RUNMC) and University Campus Biomedico, Rome (UCBM). Three different EEG systems were used to acquire the data, Brainproducts Acticaps (KCL, CIMH, RUNMC), Biosemi Active-Two (UMCU) and Micromed (UCBM). Across all sites, 70 EEG channels with standard positions were used^27^. Original sampling rates were as follows: 5000 Hz (KCL, RUNMC), 2000 Hz (CIMH), 256 Hz (UCBM) and 2048 Hz (UMCU). The recording reference was FCz. Online hardware filters were set at the manufacturer default/recommended settings. Stimuli were presented using custom-written Matlab software (KCL, CIMH, UMCU, UCBM) and Presentation (UMCU). Participants were presented with three repeated face stimuli, repeated 168 times over four blocks. Each presentation of a face could be upright or inverted (rotated 180°) with 50% of trials from each condition in pseudorandomized order. The three faces were Caucasian, African-American and Asian (from ^28^) and subtended 12.4 degrees of visual angle. Each trial began with a fixation stimulus, a colourful icon selected randomly from a set of 85 that subtended 2.9 degrees of visual angle and was positioned over where the eye region of the face would subsequently appear in both upright and inverted conditions. No icons included faces, people or other social stimuli. After a random 500-700ms interstimulus interval the face image was presented for 500ms, followed by a blank screen for 350ms. Stimuli were presented using custom-written Matlab software (KCL, CIMH, UMCU, UCBM) and Presentation (UMCU)^29^. Analyses were carried out on all facial stimuli (upright and inverted).

### Data preprocessing

#### Structural MRI data preprocessing

T1-weighted MRI images were preprocessed using a voxel-based morphometry pipeline (VBM) based on the CAT12 package (http://www.neuro.uni-jena.de/cat/) in SPM12. Images were first segmented into grey-matter, white matter and cerebrospinal fluid. Next, we created a study-specific template with DARTEL (i.e., a nonlinear diffeomorphic registration algorithm^30^) and segmented images were registered to MNI space. Flow fields from a Jacobian modulation were used to preserve the information on local tissue volume. Smoothing was performed with a 4mm full-width half-max (FWHM) isotropic Gaussian kernel.

#### Resting-state fMRI data preprocessing

After combining the three rs-fMRI scan echoes using echo-time weighted averaging, the rs-fMRI data were preprocessed using a standard preprocessing pipeline that included tools from the FMRIB Software Library (FSL version 5.0.6; http://www.fmrib.ox.ac.uk/fsl). Preprocessing included removal of the first five volumes to allow for signal equilibration, primary head motion correction via realignment to the middle volume (MCFLIRT), grand mean scaling and spatial smoothing with a 6mm FWHM Gaussian kernel. Next, we thoroughly corrected for secondary head-motion related artifacts, by applying ICA-AROMA, an ICA-based method, which automatically detects and removes motion-related components from the data^31^. ICA-AROMA has been demonstrated to remove head motion-related artifacts with high accuracy while preserving signal of interest^31,32^. Finally, we applied nuisance regression to remove signal from white matter and cerebrospinal fluid, and a high-pass filter (0.01 Hz). The rs-fMRI images of each participant were coregistered to the participants' anatomical images via boundary-based registration implemented in FSL FLIRT^33^. The T1 images of each participant were registered to MNI152 standard space using 12-parameter affine transformation and refined using non-linear registration with FSL FNIRT (10mm warp, 2mm resampling resolution). Finally, we brought all participant-level rs-fMRI images to 2mm MNI152 standard space by applying the rs-fMRI to T1 and T1 to MNI152 transformations. All further analyses were conducted in MNI152 standard space.

#### Functional task MRI data preprocessing

fMRI data analysis followed standard processing routines in SPM12 (http://www.fil.ion.ucl.ac.uk/spm/), including slice-time correction, a two-step realignment procedure, unified segmentation and normalization to standard stereotactic space as defined by the Montreal Neurological Institute (MNI), and smoothing with an 8mm full-width-at-half-maximum Gaussian Kernel. For each subject, task conditions were modelled as boxcar functions that accounted for the presentation of face blocks and shape blocks, respectively. Task regressors were convolved with the canonical hemodynamic response function (HRF) and subjected as predictors to a general linear model (GLM), along with six realignment parameters to account for head motion. During first-level model estimation, data was high-pass ﬁltered with a cut-off of 256s, and an autoregressive model of the ﬁrst order was applied. The faces condition was subsequently contrasted to the shapes condition to identify brain responses reflecting sensitivity to emotional faces.

#### EEG preprocessing

Preprocessing and harmonisation of this data was performed at Birkbeck, University of London. Each dataset was first loaded into EEGLab (Delorme & Makeig, 2004). The preprocessing pipeline is described in detail in^29^. The following steps were followed: 1) harmonisation of electrode labels to 62-channel common montage; 2) generation of horizontal electrooculogram (HEOG) channels from electrodes AF7/8 (KCL, RUNMC & UCBM only, CIMH & UMCU used external electrodes to record HEOG); 3) generation of variance-based data quality metrics and extraction of impedance values from Brainvision sites; 4) re-reference to FCz; and 5) harmonise event labels. This process resulted in harmonised data in a common EEGLab^34^ format, upon which all subsequent task-specific analyses were performed. All task processing was carried out in the Matlab Fieldtrip toolbox^35^. A bandpass filter of 0.1Hz-30Hz with 2000ms of padding to avoid filter edge-artefacts. Raw EEG data were segmented into individual trials, from -200ms to 800ms post stimulus-onset. Data were cleaned in Matlab using following criteria: a) First single trials with whole scalp artefacts and EOG artefacts (as defined in ^29^) were excluded; b) next, channels and single trials that had been interpolated previously by Mason et al.,^29^ were excluded (as interpolation is not desirable when using beamforming as described below); c) finally, individuals were dropped from analyses when less than 20 clean trials (N=9) or less than 75% of channels (N=2) per ERP were available.

### Feature extraction

#### Region of interest – fusiform gyrus

All analyses were restricted to the right and left fusiform gyrus (FFG). The FFG ROI was created by adding up four regions of the Harvard-Oxford atlas (HOA) (fMRIB, Oxford, UK) (i.e., anterior and posterior divisions of the temporal fusiform cortex, temporal occipital fusiform cortex and occipital fusiform gyrus) for both the right and left hemisphere. For rs-fMRI related analyses, we reduced the FFG ROI to have 100% coverage across all individuals’ resting-state fMRI scans. To ensure the same amount of coverage for task-fMRI data as for rs-fMRI data (without losing additional individual data), we imputed missing task-fMRI values using a scikit-learn based multivariate iterative imputer (https://scikit-learn.org/stable/modules/generated/sklearn.impute.IterativeImputer.html) within the rs-fMRI-based FFG ROIs. All T1-weighted images had 100% FFG ROI coverage, but for the sake of consistency and comparability, the structural FFG ROIs were restricted to the same sizes as for rs-fMRI and task-fMRI data. This ROI was down-sampled to larger cortical parcels in the EEG source analysis as described below.

#### Resting-state fMRI: seed-based correlation analysis

As described above, the FFG ROI was restricted to have 100% coverage across all individuals’ resting-state fMRI scans. This made up 90% of the original left FFG ROI and 94.6% of the original right FFG ROI from the HOA. To characterize the fine-grained functional subdivisions within the FFG, we conducted seed-based correlation analysis. For this, the mean time series was extracted from a spherical region of interest (6mm in diameter) centered in each participant’s peak activation voxel within the fusiform face area. This peak voxel was identified based on the individual Hariri task-fMRI T-maps within the mid-lateral FFG (mFus) and posterior lateral FFG (pFus), which correspond to the fusiform face area that are part of the probabilistic functional atlas of human occipito-temporal visual cortex area^36^. Next, Pearson’s correlation coefficient was calculated between the peak voxel time series and each voxel in the rest of the FFG before being Fisher’s z-transformed and finally smoothed with a 3D Gaussian kernel (FWHM=6mm).

#### EEG: Source analysis

Following procedures similar to those in ^37,38^, power estimates on the source level were done by applying linearly constrained minimum variance beamforming algorithm (LCMV)^39^ to the data covariance matrix and forward models of the locations of interest. The forward model was derived from the MNI ICBM 2009 template brain (<https://www.bic.mni.mcgill.ca/ServicesAtlases/ICBM152NLin2009)> using the OpenMEEG implementation^40^ of the three-layer Boundary Element Method (BEM). Both the right and left FFG ROIs were downsampled to 18 cortical parcels per left and right FFG ROI. All parcels were covered by the head model. Such parcellation scheme to a lower number of cortical parcels (vs. voxels in the other imaging modalities) was motivated by the lower spatial resolution of EEG, the spatial leakage of nearby spatial filters and the use of generic electrode positions and head models^37^. The spatial filters allow the transforming the scalp level data to source level time series at a given brain location. Subsequently, the spatial filters were multiplied with time series data resulting in source estimates time series for each cortical parcel. After applying baseline correction, the first principal component (PC) of source estimate topographies per individual across the 18 parcels per hemisphere was derived.

### Normative Modeling

Normative modelling is an emerging statistical technique that allows parsing heterogeneity by charting variation in brain-behaviour mappings relative to a normative range and provides statistical inference at the level of the individual^41^. Prior research shows that modelling cortical features as deviations from a normative neurodevelopmental trajectory and incorporating individual neurobiological variations provides more sensitive measure to parse heterogeneity^41^ and map multimodal signatures in psychopathology^42^ while also improving predictive performance^43^. Prior work addressing multimodal signatures of face processing in conduct disorder has validated this approach and shows that modelling deviations before merging different modalities is more sensitive than using raw features^42^. Normative models^41,44,45^ were trained using Bayesian Linear Regression (BLR)^46^ using PCNtoolkit package (version 0.26) (https://pcntoolkit.readthedocs.io/en/) modelling the relationship between each brain imaging modality (within the right and left FFG ROI) and age, sex and scanning site. Normative models were derived in an unbiased manner under 10-fold cross-validation. This Bayesian approach calculates the probability distribution over all functions that fit the data while specifying a prior over all possible values and relocating probabilities based on evidence (i.e., observed data). As such, it yields unbiased estimates of generalizability and inferences with increasing uncertainty with fewer data. In primary analysis, a B-spline basis expansion of the of covariate vector was used to model non-linear effects of age. We compared model performance with that of modelling age linearly (without a B-spline basis expansion). To estimate voxel-wise/time-point-wise deviations for each modality in each individual, we derived normative probability maps (NPM) that quantify the deviation from the normative model summarized in *Z* scores. These indicate the difference between the prediction (mean, $\hat{y}$_ij_) at each brain location (j) and true brain value (y_ij_) scaled by the prediction variance [expected level of variation σ^2^ij and variance learned from the normative distribution (σ^2^_nj_)]:

$$z_{ij}=\frac{y_{ij}-\hat{y}_{ij}}{\sqrt{\sigma_{ⅈj}^{2}-\sigma_{nj}^{2}}}$$

The accuracy of the normative model was evaluated using the correlation between the true and the predicted voxel values (Rho), the mean standardized log-loss, the explained variance and standardized mean squared error (Figure S1) and based on the forward models to depict the spatial / temporal representation of the voxel-wise / time pointwise normative model. For latter, we identified the peak activation within the fusiform face area based on the group t-map from the Hariri task and plotted the normative model within this peak voxel. For EEG, we plotted it at 170ms. For all modalities this was done in males and in the largest acquisition site (KCL). Figure S3 depicts these normative models per modality.

Finally, to assess whether autistic and non-autistic individuals differed in their extreme deviations, the NPMs were thresholded at an absolute value of Z>|2.6| (i.e., *p* < 0.005)^42,47–49^. Based on this fixed threshold, we defined extreme positive and extreme negative deviations for each participant. All extreme deviations per individual were summarized into scores representing the percentage of extreme positive and extreme negative deviations per individual in relation to the total number of voxels. These percentage scores were compared between autistic and non-autistic individuals using a non-parametric Mann-Whitney U-test.

### Linked Independent Component Analysis

Linked Independent Component Analysis (LICA)^24,42,50–54^ is a Bayesian tensor extension of single modality ICA model which provides an automatic decomposition of the brain features into independent components (ICs) that characterize the inter-subject brain variability. These multiple tensor decompositions share a mixing matrix or subject course across individual feature factorizations that reflect the subject contributions to each independent component. These subject loadings per IC can be used to investigate the relationship between the brain phenotypes and demographic and clinical measures. Further, each IC also provides a map of spatial variation per modality and a vector reflecting the relative contribution of each modality to the component. The advantage of LICA in comparison to other multimodal methods is that each modality can have completely different numbers of features, spatial correlations, intensity distributions and units, given that LICA optimally weighs the contributions of each modality by the correction for the number of effective degrees of freedom and the use of automatic relevance determination (ARD) priors on components. Also, non-Gaussian spatial sources are more likely to represent actual structured signals in the data^50,52,55^.

Here, we used LICA to merge the unthresholded Z-deviation maps across the four different imaging modalities within the right and left FFG ROIs. Each measure per hemisphere was treated as a different ‘modality’ (right structure, left structure, right rs-fMRI, left rs-fMRI, right task-fMRI, left task-fMRI, right EEG, left EEG) resulting in eight input maps (modalities). Hemispheres were treated separately to study the hemispheric contributions and model the different noise characteristics individually. Based on our sample size and following recommendations described in earlier papers^24,42,51,53,54^ (i.e., sample size N / 4), we estimated 50 independent components. We additionally calculated a multimodal index per independent component to quantify each IC’s multimodal nature as previously described in ^53^:

$$Multimodality Index=1-n*\frac{score(IC)}{n-1}$$

Where:

$$score(IC)=max(weight\left( IC \right))-\frac{1}{n}$$

n = number of modalities

### Characterization of Spatial Maps

The spatial maps were converted to pseudo-Z-statistics taking into account the scaling of the variables and the SNR in each modality. To characterize the spatial maps of significantly implicated ICs both anatomically and functionally, the spatial Z-maps of each imaging modality were thresholded at the 95^th^ percentile and next, the percentage of overlap of suprathreshold voxels with both an anatomical and functional atlas was quantified. The four subregions of the fusiform gyrus derived from the Harvard-Oxford atlas (HOA) (fMRIB, Oxford, UK) (i.e., anterior and posterior divisions of the temporal fusiform cortex, temporal occipital fusiform cortex and occipital fusiform gyrus) were selected for anatomical characterisation. The different subregions covering the FFG derived from the probabilistic functional atlas of human occipito-temporal visual cortex (VIS-atlas)^36^ of early visual and category-selective regions (i.e., faces, bodies, characters, places) were used for functional characterisation.

### Support Vector Machine

To assess whether multimodal components outperformed unimodal components in discriminating autistic from non-autistic individuals, we implemented a Support Vector Machine (SVM) classification algorithm under 10-fold cross-validation using scikit-learn in Python. Features were either the unimodal or multimodal components. A threshold of 90% was selected to define multimodality, where no single imaging modality (regardless of hemisphere) contributed more than 90% to each component. The remaining components were labelled as unimodal. We used the default linear kernel and penalty parameter of 1, along with class-weighting to account for group size differences between autistic and non-autistic individuals.

The area under the receiver operating characteristic curve (AUC) was used as the performance metric to assess the classifier's discrimination ability. To test for significant differences in AUC between multimodal and unimodal components, we generated a null distribution of AUC differences by shuffling the cross-validated scores 10.000 times and re-evaluating the classifier performance and computed the likelihood of observing the observed AUC difference under the null hypothesis. To test for robustness of results across different multimodal thresholds, we ran sensitivity analyses across different thresholds resulting in slightly varying degrees of multimodality ranging between 85% to 99% of single modality contributions. Given that each threshold resulted in a different number of unimodal vs. multimodal components (e.g., threshold of 90% resulted in 11 multimodal vs. 39 unimodal ICs), we further checked whether results remained stable when forcing uni- and multimodal components to have the same amount of features. For this we selected the top most multimodal and top most unimodal components varying between one to 22 (corresponding to 98% of multimodality contribution) numbers of ICs.

### Canonical Correlation Analysis

To model the multivariate relationship between the identified multimodal ICs and cognitive features related to social functioning and face processing in autism, we ran canonical correlation analysis (CCA)^56^. CCA is a multivariate approach to simultaneously model two sets of linear projections (based on the brain-related independent components and the cognitive features) and maximizes their correlation. Cognitive features were z-standardized. We controlled for age, sex and scanning site using the Huh-Jhun residualization method as described in by Winkler et al^57^. The statistical significance of the CCA modes was assessed by a complete permutation inference algorithm, where both brain and behaviour data were permuted separately across all participants with 10,000 iterations^57^. We corrected for multiple comparisons of each CCA mode using a stepwise cumulative maximum approach (p<0.05) as described in Winkler et al.^57^. Canonical variates were calculated respectively for the brain and behavioural sets by multiplying the canonical coefficients with the original sets. Each pair of canonical variates is one CCA mode. The brain-behaviour relationship was evaluated based on how much each multimodal IC and how much each cognitive feature contributed to the correlation. These contributions were measured by the respective loadings which are based on Haufe-transformed canonical coefficients^58^.

In order to assess robustness of CCA results, as previously, we set a range of multimodal thresholds between 85% to 99% and selected components with modality contributions exceeding this threshold as multimodal. We then re-ran the CCA for each threshold to assess stability of results across slightly varying degrees of multimodality.

### References

1.	Loth, E. *et al.* The EU-AIMS Longitudinal European Autism Project (LEAP): Design and methodologies to identify and validate stratification biomarkers for autism spectrum disorders. *Molecular Autism* **8**, (2017).

2.	Charman, T. *et al.* The EU-AIMS Longitudinal European Autism Project (LEAP): Clinical characterisation. *Molecular Autism* **8**, (2017).

3.	Del Barrio, V. Diagnostic and statistical manual of mental disorders. in *The Curated Reference Collection in Neuroscience and Biobehavioral Psychology* (2016). doi:10.1016/B978-0-12-809324-5.05530-9.

4.	American Psychiatric Association. *Diagnostic and statistical manual of mental disorders: DSM-IV-TR (text revision)*. *American Journal of Psychiatry* (2000).

5.	American Psychiatric Association. *Diagnostic and Statistical Manual of Mental Disorders 1. American Psychiatric Association.* *Arlington* (2013). doi:10.1176/appi.books.9780890425596.893619.

6.	World Health Organization. *The ICD-10 classification of mental and behavioural disorders: Diagnostic criteria for research*. *The ICD-10 classification of mental and behavioural disorders: Diagnostic criteria for research* (1993).

7.	Lord, C., Rutter, M. & Le Couteur, A. Autism Diagnostic Interview-Revised: a revised version of a diagnostic interview for caregivers of individuals with possible pervasive developmental disorders. *Journal of autism and developmental disorders* **24**, 659–685 (1994).

8.	Lord, C. *et al.* The autism diagnostic observation schedule-generic: a standard measure of social and communication deficits associated with the spectrum of autism. *Journal of autism and developmental disorders* **30**, 205–223 (2000).

9.	Wechsler, D. *Wechsler Abbreviated Scale of Intelligence–Second Edition (WASI‐II).* (NCS Pearson., 2011).

10.	Wechsler, D. *Wechsler Intelligence Scale for Children (3rd ed.)*. (Psychological Corporation, 1991).

11.	Wechsler, D. *Wechsler Intelligence Scale for Children (4th ed.)*. (Psychological Corporation, 2003).

12.	Wechsler, D. *Wechsler Adult Intelligence Scale (3rd ed.)*. (The Psychological Corporation, 1997).

13.	Wechsler, D. *Wechsler Adult Intelligence Scale–Fourth Edition*. (Pearson, 2008).

14.	Hus, V., Gotham, K. & Lord, C. Standardizing ADOS domain scores: Separating severity of social affect and restricted and repetitive behaviors. *Journal of Autism and Developmental Disorders* **44**, 2400–2412 (2014).

15.	Gotham, K., Risi, S., Pickles, A. & Lord, C. The autism diagnostic observation schedule: Revised algorithms for improved diagnostic validity. *Journal of Autism and Developmental Disorders* **37**, (2007).

16.	Hariri, A. R., Tessitore, A., Mattay, V. S., Fera, F. & Weinberger, D. R. The amygdala response to emotional stimuli: A comparison of faces and scenes. *NeuroImage* **17**, (2002).

17.	Baron-Cohen, S., Wheelwright, S., Hill, J., Raste, Y. & Plumb, I. The ‘Reading the Mind in the Eyes’ Test revised version: A study with normal adults, and adults with Asperger syndrome or high-functioning autism. *Journal of Child Psychology and Psychiatry and Allied Disciplines* **42**, (2001).

18.	Auyeung, B. *et al.* The children’s empathy quotient and systemizing quotient: Sex differences in typical development and in autism spectrum conditions. *Journal of Autism and Developmental Disorders* **39**, (2009).

19.	Auyeung, B., Allison, C., Wheelwright, S. & Baron-Cohen, S. Brief report: Development of the adolescent empathy and systemizing quotients. *Journal of Autism and Developmental Disorders* **42**, (2012).

20.	Baron-Cohen, S., Richler, J., Bisarya, D., Gurunathan, N. & Wheelwright, S. The systemizing quotient: An investigation of adults with Asperger syndrome or high-functioning autism, and normal sex differences. *Philosophical Transactions of the Royal Society B: Biological Sciences* **358**, (2003).

21.	Bodfish, J. W., Symons, F. J., Parker, D. E. & Lewis, M. H. Repetitive Behavior Scale–Revised. *PsycTESTS®* (2000).

22.	Tomchek, S. D. & Dunn, W. Sensory processing in children with and without autism: a comparative study using the short sensory profile. *American Journal of Occupational Therapy* **61**, (2007).

23.	Llera, A. *et al.* Evaluation of data imputation strategies in complex, deeply-phenotyped data sets: the case of the EU-AIMS Longitudinal European Autism Project. *BMC Medical Research Methodology* **22**, 229 (2022).

24.	Mei, T. *et al.* Autism Is Associated With Interindividual Variations of Gray and White Matter Morphology. *Biol Psychiatry Cogn Neurosci Neuroimaging* S2451-9022(22)00212–9 (2022) doi:10.1016/j.bpsc.2022.08.011.

25.	Looden, T. *et al.* Patterns of connectome variability in autism across five functional activation tasks: findings from the LEAP project. *Molecular Autism* **13**, 53 (2022).

26.	Kundu, P., Inati, S. J., Evans, J. W., Luh, W. M. & Bandettini, P. A. Differentiating BOLD and non-BOLD signals in fMRI time series using multi-echo EPI. *NeuroImage* **60**, (2012).

27.	Oostenveld, R. & Praamstra, P. The five percent electrode system for high-resolution EEG and ERP measurements. *Clin Neurophysiol* **112**, 713–719 (2001).

28.	Tye, C. *et al.* Neurophysiological responses to faces and gaze direction differentiate children with ASD, ADHD and ASD+ADHD. *Dev Cogn Neurosci* **5**, 71–85 (2013).

29.	Mason, L. *et al.* Stratifying the autistic phenotype using electrophysiological indices of social perception. *Science Translational Medicine* **14**, eabf8987 (2022).

30.	Ashburner, J. A fast diffeomorphic image registration algorithm. *NeuroImage* **38**, 95–113 (2007).

31.	Pruim, R. H. R. *et al.* ICA-AROMA: A robust ICA-based strategy for removing motion artifacts from fMRI data. *NeuroImage* **112**, 267–277 (2015).

32.	Parkes, L., Fulcher, B., Yücel, M. & Fornito, A. An evaluation of the efficacy, reliability, and sensitivity of motion correction strategies for resting-state functional MRI. *NeuroImage* **171**, 415–436 (2018).

33.	Greve, D. N. & Fischl, B. Accurate and robust brain image alignment using boundary-based registration. *NeuroImage* **48**, 63–72 (2009).

34.	Delorme, A. & Makeig, S. EEGLAB: an open source toolbox for analysis of single-trial EEG dynamics including independent component analysis. *J Neurosci Methods* **134**, 9–21 (2004).

35.	Oostenveld, R., Fries, P., Maris, E. & Schoffelen, J. M. FieldTrip: Open source software for advanced analysis of MEG, EEG, and invasive electrophysiological data. *Computational Intelligence and Neuroscience* **2011**, (2011).

36.	Rosenke, M., van Hoof, R., van den Hurk, J., Grill-Spector, K. & Goebel, R. A Probabilistic Functional Atlas of Human Occipito-Temporal Visual Cortex. *Cerebral Cortex* **31**, 603–619 (2021).

37.	Popov, T. *et al.* Test–retest reliability of resting-state EEG in young and older adults. *Psychophysiology* **60**, e14268 (2023).

38.	Mahjoory, K. *et al.* Consistency of EEG source localization and connectivity estimates. *NeuroImage* **152**, 590–601 (2017).

39.	Van Veen, B. D., van Drongelen, W., Yuchtman, M. & Suzuki, A. Localization of brain electrical activity via linearly constrained minimum variance spatial filtering. *IEEE Trans Biomed Eng* **44**, 867–880 (1997).

40.	Gramfort, A., Papadopoulo, T., Olivi, E. & Clerc, M. OpenMEEG: opensource software for quasistatic bioelectromagnetics. *BioMedical Engineering OnLine* **9**, 45 (2010).

41.	Marquand, A. F. *et al.* Conceptualizing mental disorders as deviations from normative functioning. *Molecular Psychiatry* **24**, 1415–1424 (2019).

57.	Winkler, A. M., Renaud, O., Smith, S. M. & Nichols, T. E. Permutation inference for canonical correlation analysis. *NeuroImage* **220**, (2020).

58.	Haufe, S. *et al.* On the interpretation of weight vectors of linear models in multivariate neuroimaging. *NeuroImage* **87**, 96–110 (2014).
