## Supplementary Figures for "A multimodal neural signature of face processing in autism within the fusiform gyrus"

**Figure S1**

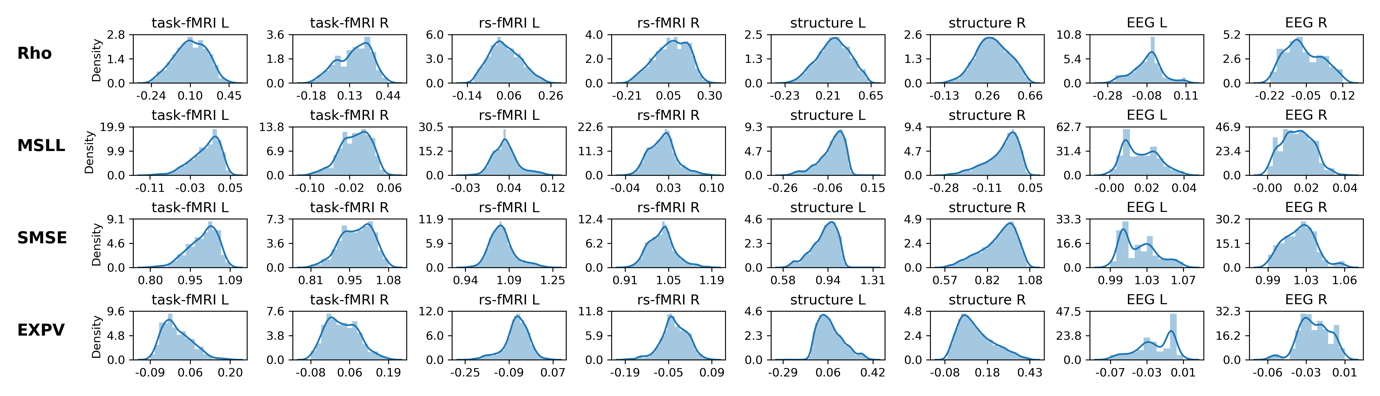

**Figure S1.** Evaluation of normative model performance for each imaging modality per hemisphere when using BLR with B-Spline to model non-linear age effects. The upper row shows the coorelation between the true and predicted values (Rho; closer to 1 = better model performance), the second row shows the mean standardized log-loss (MSLL; more negative = better model performance), the third row shows the standardized mean squared error (SMSE; closer to 0 = better model performance) and the last row shows the explained variance (EXPV; closer to 1 = better model performance). Abberviations: MSLL= mean standardized log-loss, SMSE=standarized mean squared error, EXPV=explained variance.

**Figure S2**

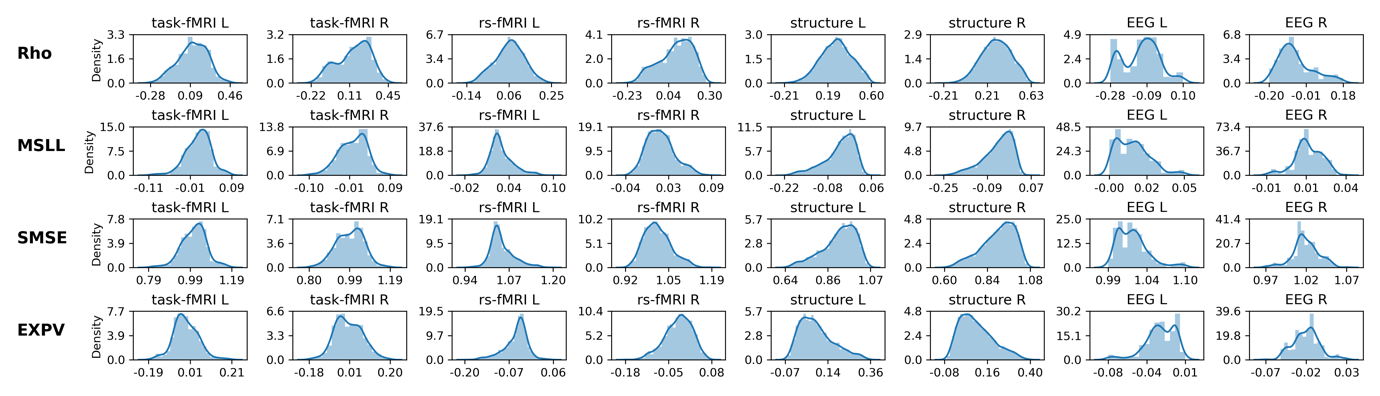

**Figure S2.** Evaluation of normative model performance for each imaging modality per hemisphere when using BLR without a B-Spline basis extension to model linear age effects. Evaluations metrics are highly similar. The upper row shows the coorelation between the true and predicted values (Rho; closer to 1 = better model performance), the second row shows the mean standardized log-loss (MSLL; more negative = better model performance), the third row shows the standardized mean squared error (SMSE; closer to 0 = better model performance) and the last row shows the explained variance (EXPV; closer to 1 = better model performance). Abberviations: MSLL= mean standardized log-loss, SMSE=standarized mean squared error, EXPV=explained variance.

**Figure S3**

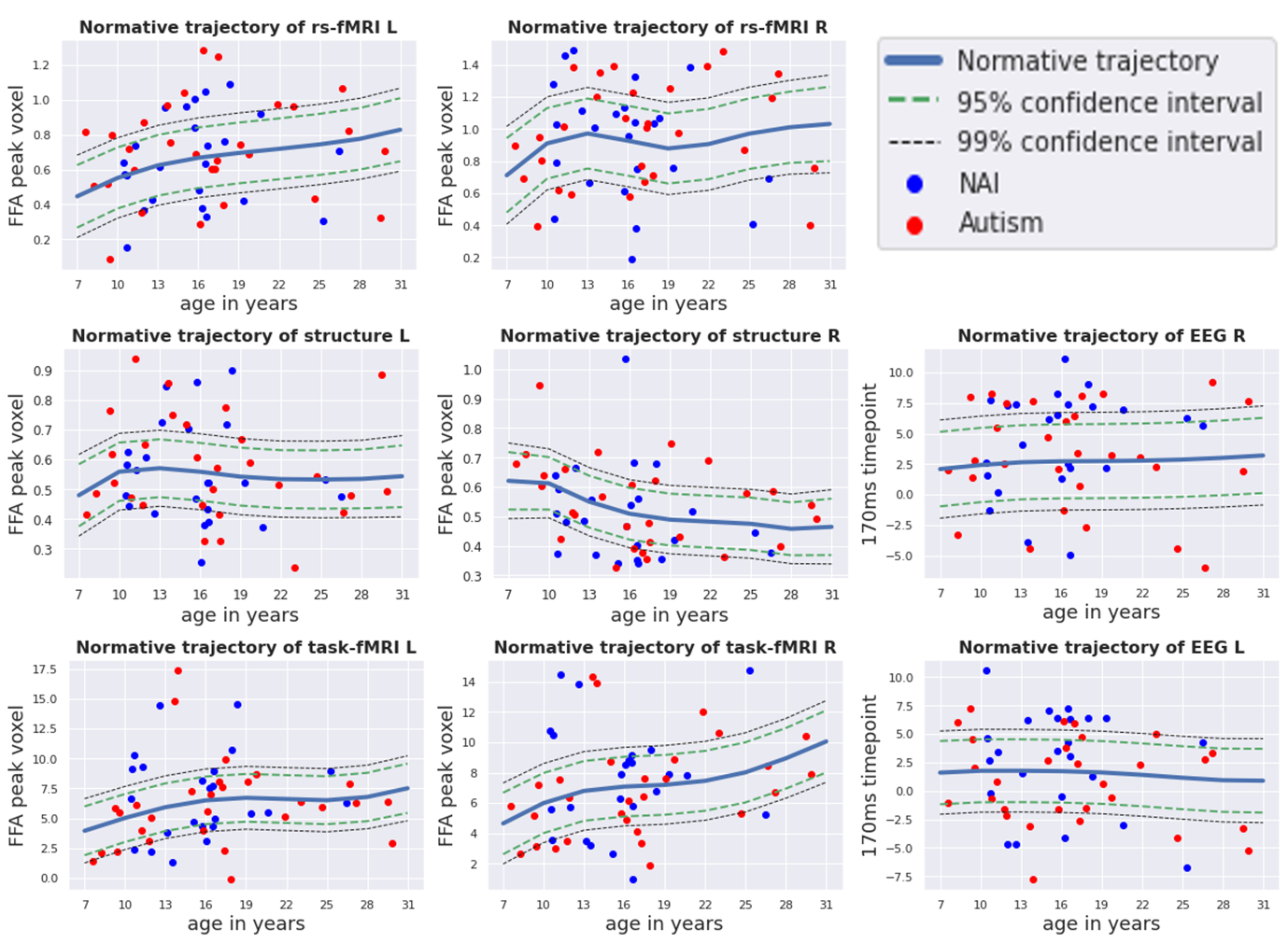

**Figure S3**. The forward models depicting the spatial and temporal representations of the voxel-wise / time pointwise normative model for each imaging modality per hemisphere. For imaging modalities (structure, task-fMRI, resting-state fMRI) the normative model of the peak activation voxel within the fusiform face area is shown, whereas for EEG at timepoint 170ms. The regression line depicts the predicted values between 7 and 30 years of age along with centiles of confidence (95^th^ and 99^th^). The blue dots are the true values for non-autistic males in the KCL site, whereas the red dots are the true values for the autistic males in the KCL site.

**Figure S4**

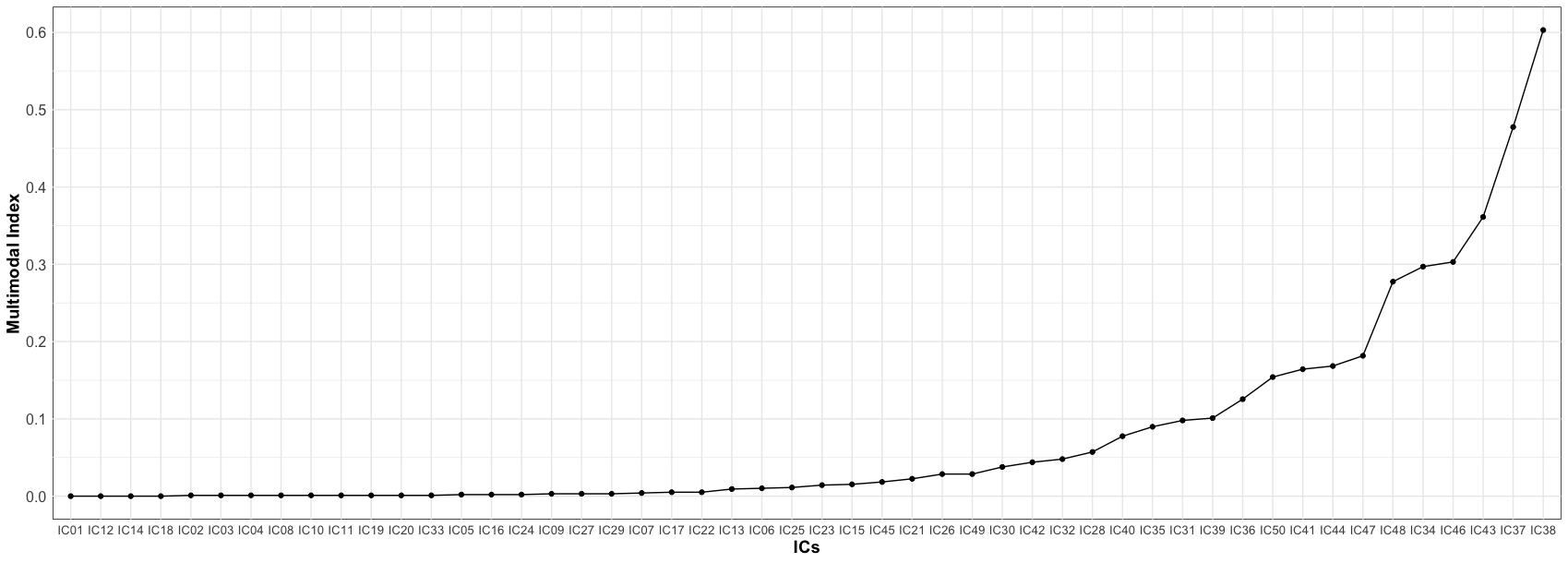

**Figure S4.** The multimodal Index ranging between 0 (dominance by one single modality) to (equal contribution of all modalities) is shown as a function of all 50 ICs. In order to pick the most multimodal components, we used a cut-off of 0.1 which also corresponded to no single modality contributing more than 90% to each independent component.

**Figure S5**

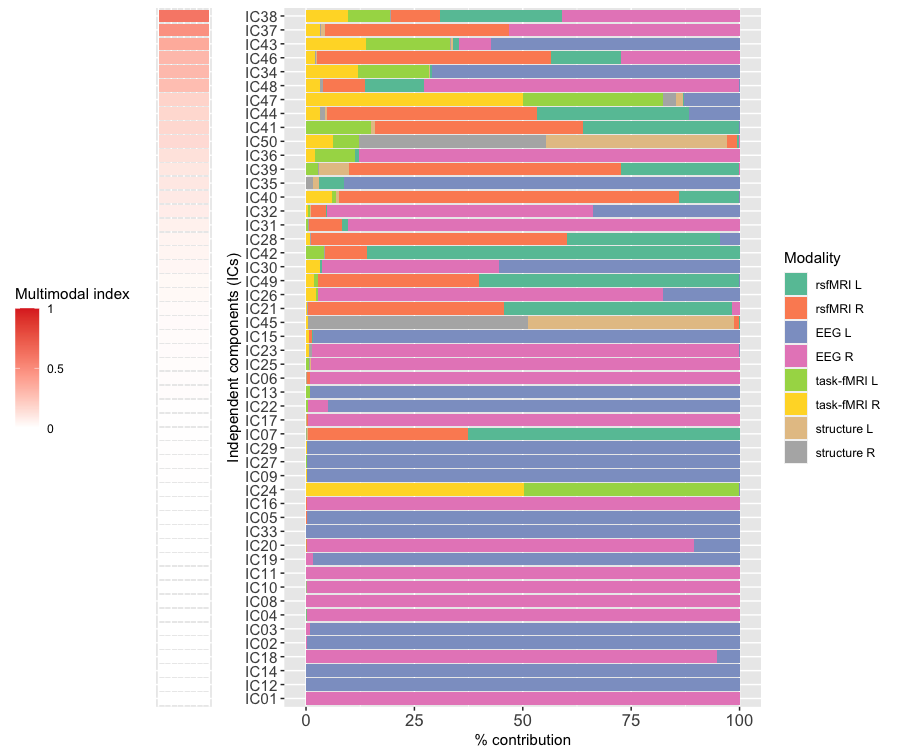

**Figure S5.** We identified 50 independent components using Linked ICA. These are ordered according to how multimodal they are (i.e., multimodal index). Each color represents one of the eight feature maps (‘modalities’) fed into the model. Overall, across these, the right hemisphere (51.7%) and the left hemisphere (48.3%) showed equal contributions. Single modality contributions were as follows: EEG R (35.0%) > EEG L (33.2%) > rs-fMRI L (11.2%) > rs-fMRI R (9.6%) > task-fMRI R (3.5%) > task-fMRI L (3.4%) > structure L (2.1%) > structure R (2.1%).

**Figure S6**

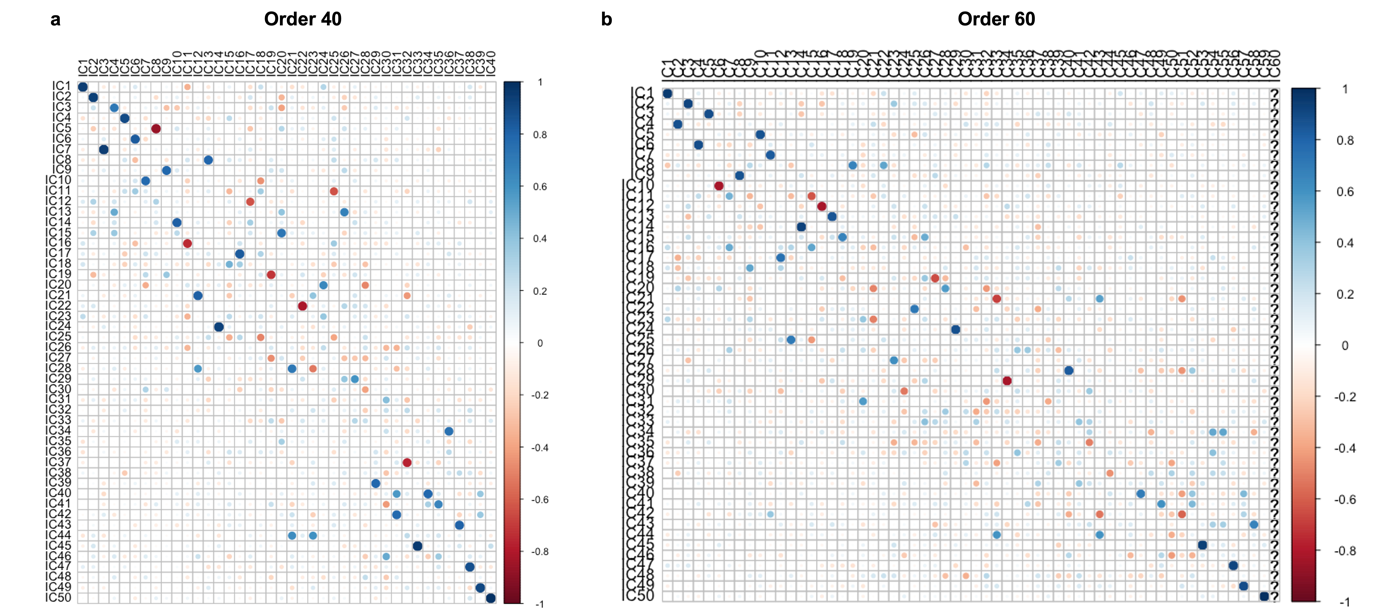

**Figure S6**. To evaluate the robustness of our selected model order (N=50), we re-ran LICA using different dimensional factorization of subject loadings (N=40 and N=60) and correlated the reported (N=50) dimensional factorization subject loadings with the alternative dimensional factorizations. Most components were recovered with high accuracy independently of the order of the factorization.

**Figure S7**

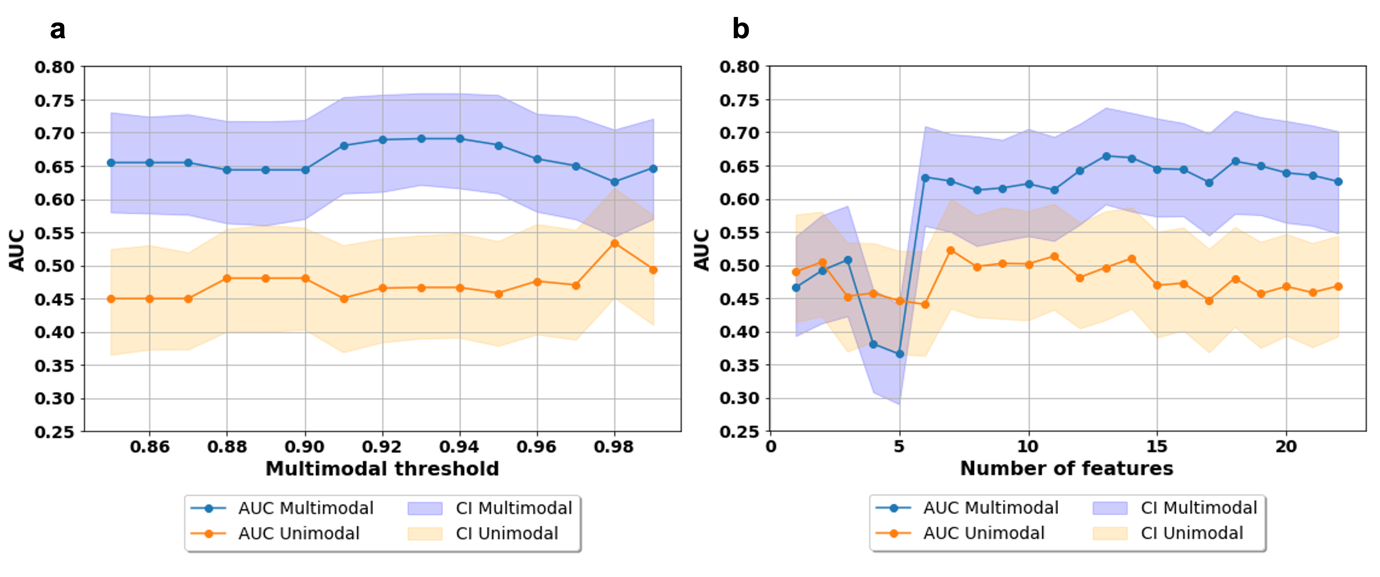

**Figure S7.** We ran a support vector machine (SVM) classification algorithm to test whether the multimodal ICs outperformed the unimodal ICs in discriminating autistic from non-autistic individuals. The graph in Figure S7a shows the area under the receiver operating characteristic curve (AUC) along with the 95% confidence interval (CI) as a function of different thresholds between 85% to 99% that define whether a component is multimodal or unimodal. Graph in Figure S7b shows the AUC when forcing uni- and multimodal features to have the same amount of ICs in each fold. In the beginning (up to six ICs) there are no differences, which become apparent when increasing the number of ICs included as features.

**Figure S8**

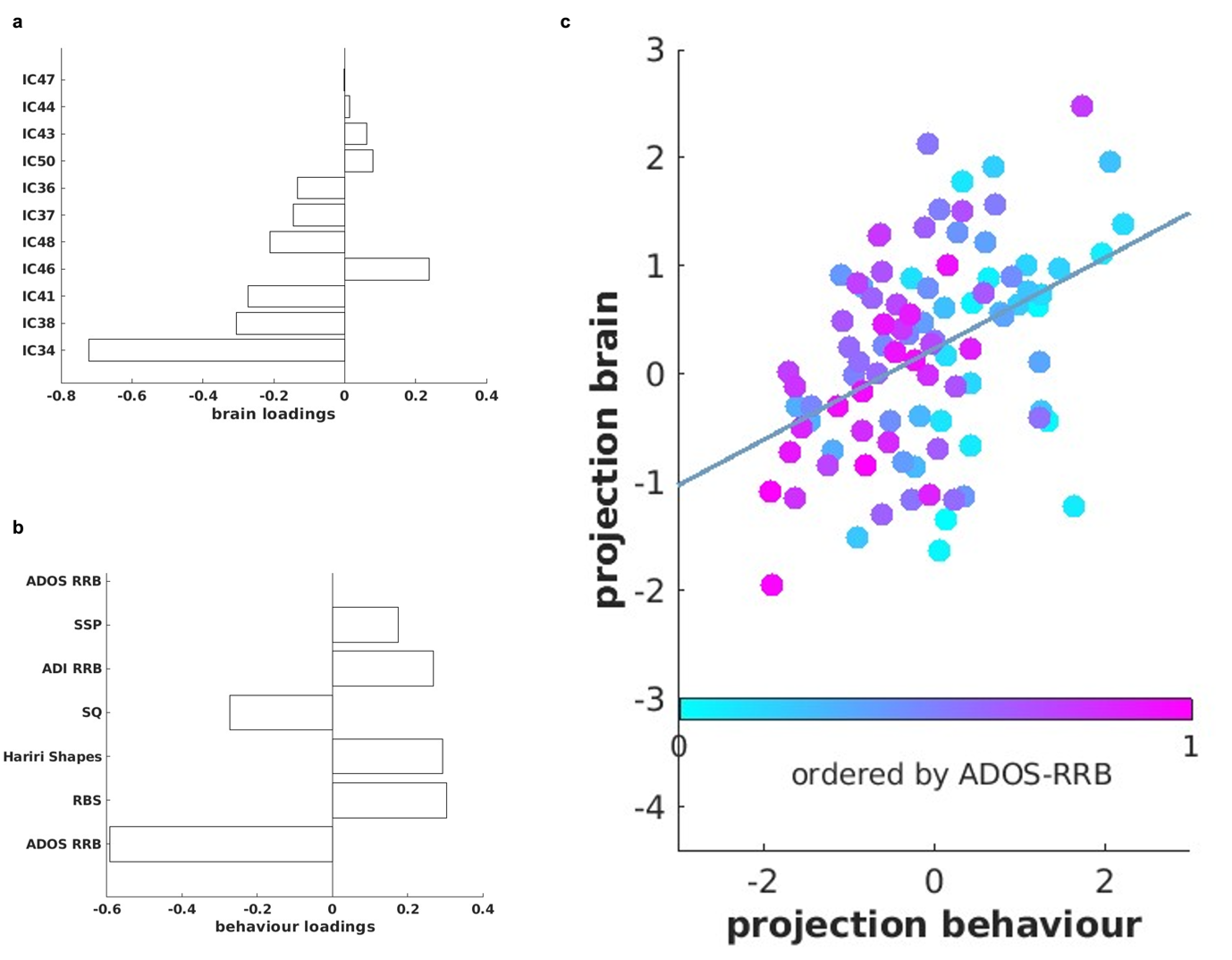

**Figure S8.** The multivariate association (i.e., canonical correlation) was not significant between the eleven multimodal ICs and the non-social features associated with autism (*r*=0.49, *p*_FDR_=0.51). Figure S8a shows the loadings of each multimodal component contributing to the CCA mode, while Figure S8b shows the loadings of each non-social feature contributing to the CCA mode. Figure S8c shows the canonical correlation scatterplot color-coded by the highest contributing non-social feature (ADOS RRB). The x-axis depicts the projected behavioural CCA variate and the y-axis the multimodal ICs CCA variates.

**Figure S9**

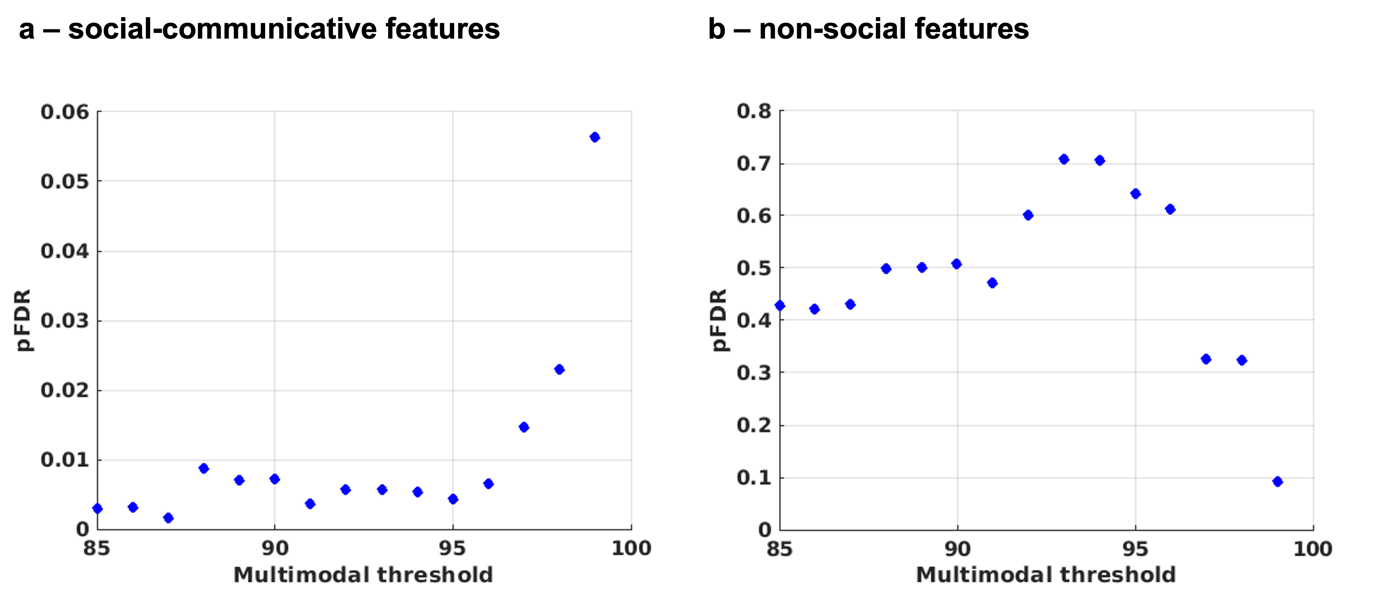

**Figure S9.** We ran two separate canonical correlation analyses (CCA) to model the relationship between multimodal independent components and either a) social-communicative features related to autism or b) non-social features related to autism (e.g., restricted repetitive behaviours, sensory processing). We tested robustness of the CCA across slightly varying degrees of multimodality by changing the multimodality threshold ranging between 85%-99% (by steps of 1%). Results for both associations remained stable.

**Figure S10**

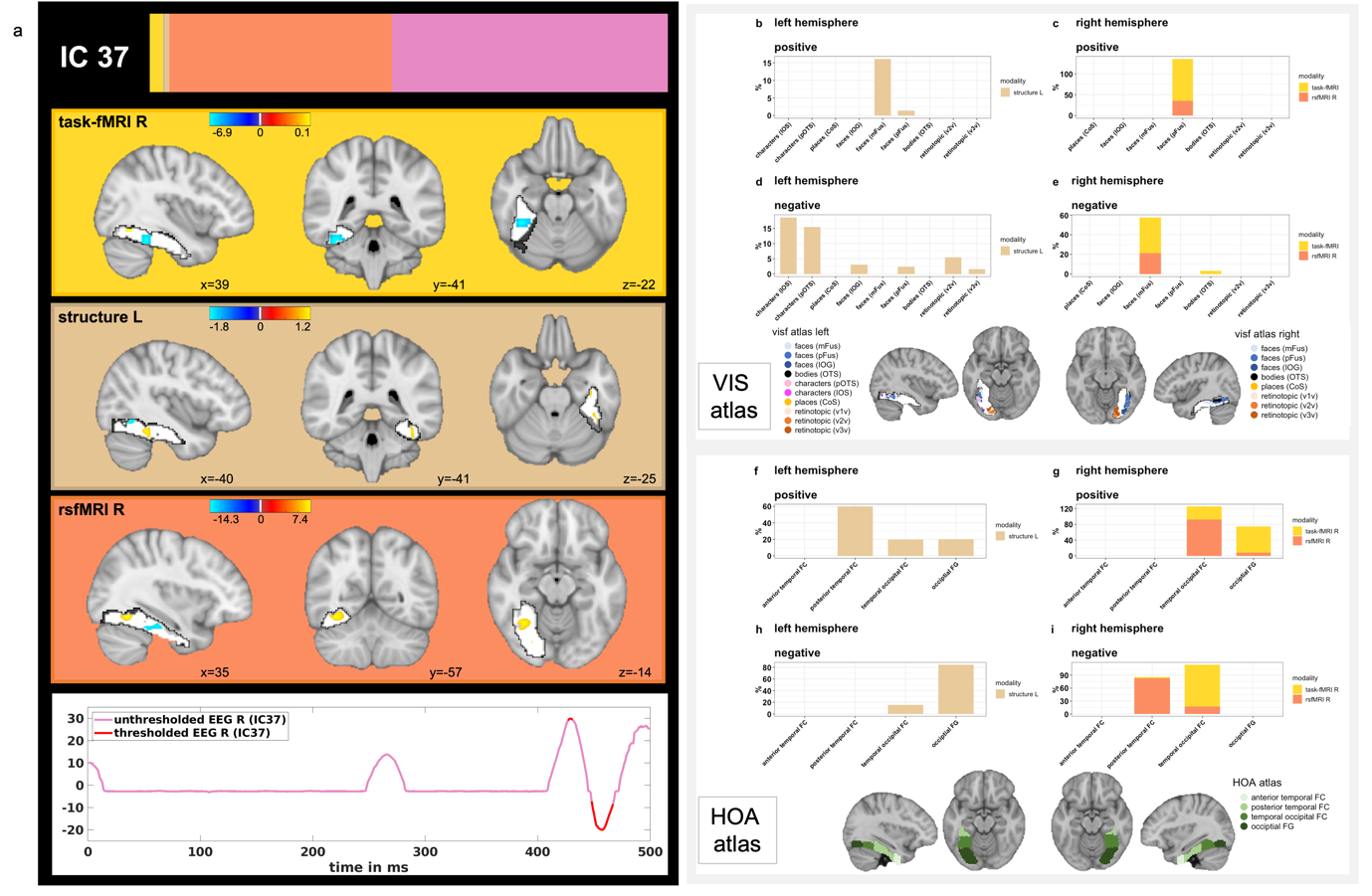

**Figure S10**. Spatial and temporal patterns of the different neuroimaging modalities associated with IC37. Figure S10a shows the spatial and temporal Z-maps thresholded at the 95th percentile. Figures S10b-e depict the spatial overlap of suprathreshold voxels with a probabilistic functional atlas of the occipito-temporal cortex (i.e., VIS-atlas). Figures S10f-i show the spatial overlap of suprathreshold voxels with the structural Harvard-Oxford atlas and the four subregions of the fusiform gyrus (i.e., anterior and posterior divisions of the temporal fusiform cortex, temporal occipital fusiform cortex and occipital fusiform gyrus). Here, Figures S10b/f and S10c/g show the positive loadings and Figures S10d/h and S10e/i the negative loadings, whereas Figures S10b/f and S10d/h depict the left hemisphere and Figures S10c/g and S10e/i the right hemisphere.

**Figure S11**

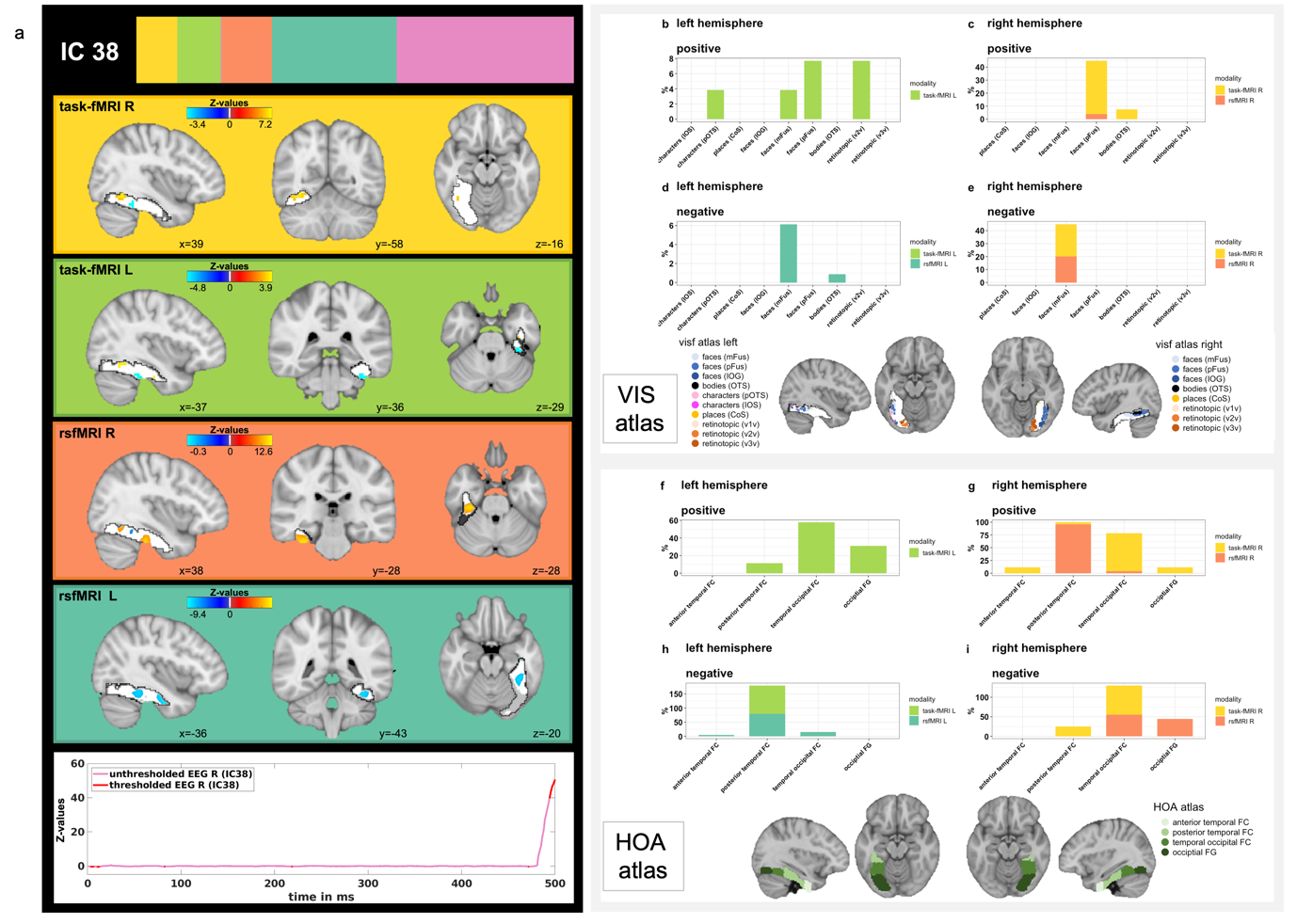

**Figure S11**. Spatial and temporal patterns of the different neuroimaging modalities associated with IC38. Figure S11a shows the spatial and temporal Z-maps thresholded at the 95th percentile. Figures S11b-e depict the spatial overlap of suprathreshold voxels with a probabilistic functional atlas of the occipito-temporal cortex (i.e., VIS-atlas). Figures S11f-i show the spatial overlap of suprathreshold voxels with the structural Harvard-Oxford atlas and the four subregions of the fusiform gyrus (i.e., anterior and posterior divisions of the temporal fusiform cortex, temporal occipital fusiform cortex and occipital fusiform gyrus). Here, Figures S11b/f and S11c/g show the positive loadings and Figures S11d/h and S11e/i the negative loadings, whereas Figures S11b/f and S11d/h depict the left hemisphere and Figures S11c/g and S11e/i the right hemisphere.

**Figure S12**

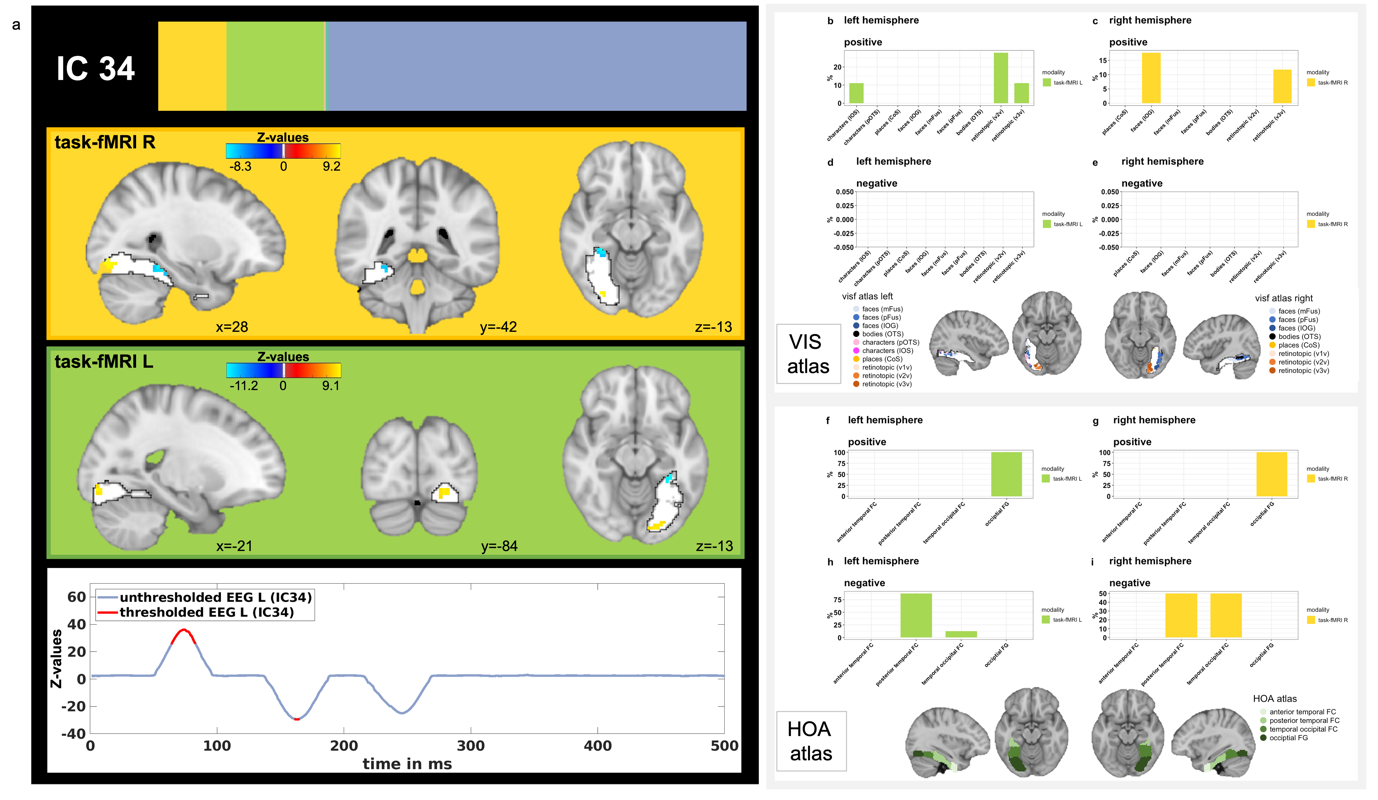

**Figure S12**. Spatial and temporal patterns of the different neuroimaging modalities associated with IC38. Figure S12a shows the spatial and temporal Z-maps thresholded at the 95th percentile. Figures S12b-e depict the spatial overlap of suprathreshold voxels with a probabilistic functional atlas of the occipito-temporal cortex (i.e., VIS-atlas). Figures S12f-i show the spatial overlap of suprathreshold voxels with the structural Harvard-Oxford atlas and the four subregions of the fusiform gyrus (i.e., anterior and posterior divisions of the temporal fusiform cortex, temporal occipital fusiform cortex and occipital fusiform gyrus). Here, Figures S12b/f and S12c/g show the positive loadings and Figures S12d/h and S12e/i the negative loadings, whereas Figures S12b/f and S12d/h depict the left hemisphere and Figures S12c/g and S12e/i the right hemisphere.

**Figure S13**

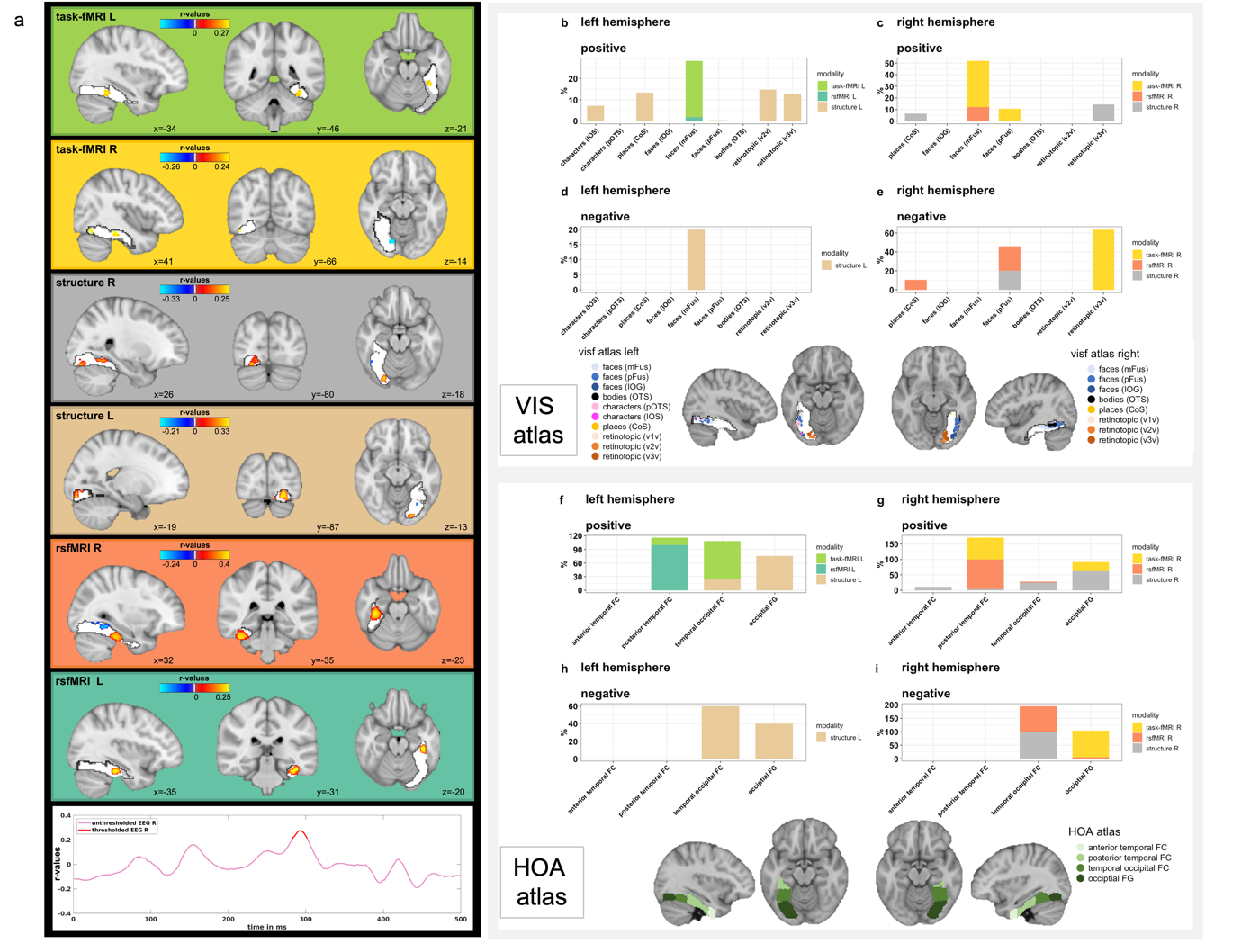

**Figure S13.** To visualize the spatial and temporal patterns of each imaging modality associated with each clinical cognitive measure, we computed the correlations between the original imaging data (i.e., the Z-deviations maps) and the canonical imaging variate derived from the CCA. Significance of correlation maps was assessed with 1000 permutations and significantly positively (yellow) or negatively (blue) correlated clusters / timepoints are visualized in Figure S13a. Figures S13b-e depict the spatial overlap of significant voxels with a probabilistic functional atlas of the occipito-temporal cortex (i.e., VIS-atlas). Figures S13f-i show the spatial overlap of significant voxels with the structural Harvard-Oxford atlas and the four subregions of the fusiform gyrus (i.e., anterior and posterior divisions of the temporal fusiform cortex, temporal occipital fusiform cortex and occipital fusiform gyrus). Here, Figures S13b/f and S13c/g show the positive associations and Figures S13d/h and S13e/i the negative associations, whereas Figures S13b/f and S13d/h depict the left hemisphere and Figures S13c/g and S13e/i the right hemisphere.
